# Low Genetic Risk for Coronary Artery Disease underlies Multigenerational Longevity and Healthy Aging

**DOI:** 10.64898/2025.12.03.25341464

**Authors:** Pedro Sant’Anna Barbosa Ferreira, Stella Trompet, P. Eline Slagboom, Joris Deelen, Marian Beekman, Niels van den Berg

## Abstract

Descendants of exceptionally long-lived families have a delayed onset of their first chronic disease. We therefore hypothesize that one of the key features explaining healthy survival up to high ages is the absence of chronic disease risk alleles. Using the LLS and Leiden85+ multigenerational cohorts and Polygenic Scores (PGS) for chronic diseases, we showed that an increasing number of long-lived ancestors is additively associated with lower genetic risk for coronary artery disease (CAD). We further showed that a lower PGS for CAD explains up to 20% of the delay in cardiovascular disease incidence in long-lived families. Finally, we constructed a novel cholesterol-metabolism-PGS, based on gene-annotation enrichment analysis, that predicted time to all-cause mortality in two 90+ study populations. Our findings demonstrate that the absence of chronic disease risk alleles is one key-feature linked to longevity and that alleles linked to cholesterol metabolism are a key component in healthy aging trajectories.

## Introduction

Aging is the largest risk factor for functional impairments and chronic diseases^1^, including cardiovascular conditions, which are the leading cause of death worldwide^2^. With advancing age, the risk of developing multiple health conditions (multimorbidity) rises. By the age of 65, around 70% of the people have at least one chronic disease, with an average of four diseases per person^3^. Given that the proportion of older adults in the population is steadily rising^4^, the already substantial burden of multimorbidity is expected to increase further^5^, placing growing pressure on healthcare systems and highlighting the urgent need for novel strategies to promote healthy aging^6–8^. Yet, some individuals defy this general trend by reaching exceptionally high ages while delaying or even avoiding the onset of chronic diseases^9,10^. Remarkably, this longevity trait clusters within families, indicating that the presence of protective genetic variants, the absence of deleterious variants, and shared environmental influences may all play a role. Thus, studying these exceptionally old individuals and their family members may provide valuable insights into the pathways of healthy aging, the markers and mechanisms of resilience, and vulnerability in older age, which is key for providing adequate clinical care ^11,12^.

Healthy survival into advanced age, a key feature of longevity, suggests that the absence or delayed onset of chronic diseases is an important characteristic underlying this trait. Since chronic diseases are partially explained by genetic factors, it has been hypothesized that the absence of alleles that predispose individuals to these conditions is one of the key-features in achieving longevity. Previous research has investigated this hypothesis without favourable evidence^13–16^. One factor that could explain the results from previous studies is the limited power to observe marked absence of genetic determinants due to small sample sizes and number of alleles identified at the time. Another factor that limited the success of genetic longevity studies is the heterogeneity in the criteria used to identify individuals carrying the heritable component of longevity^17–19^. Recent research showed that longevity is transmitted as a quantitative trait when persons belong to the oldest 10% survivors of their own birth cohort and if at least 30% of their ancestors also belong to the oldest 10% survivors of their respective birth cohort^20^. Given the improvements over the last century in nutrition, lifestyle, and healthcare, there are individuals who may reach exceptional old ages without carrying the heritable component of longevity (likely representing phenocopies)^21^. Hence, incorporating an individual’s familial history for longevity is a key component in reducing phenotypic heterogeneity when selecting participants for longevity studies and thereby avoiding phenocopies.

The past decade has seen a strong increase in the power of genome-wide association studies (GWASs) resulting in the discovery of numerous additional disease-associated alleles for age-related diseases^22^. Benefiting from these more powered GWASs, recent studies^23–25^ revisited the hypothesis that exceptionally old people carry fewer disease risk alleles than the general population. Using Polygenic Scores (PGSs) to quantify the genetic propensity to a disease and employing no systematic approach for the selection of which diseases to test, these studies obtained mixed results. Furthermore, a deeper mechanistic understanding of the results is desired to identify which genetic risk avoidance contributes to healthy survival up to high ages. We identified four key issues in the literature where further investigation is needed to unravel the genetic basis of human longevity through a focus on absence of disease risk alleles: a) a systematic approach where the selection of diseases is based on the most common causes of death within the background population has yet to be employed, b) research has not yet shown if having an increasing number of exceptional old ancestors associates with a reduced number of disease risk alleles, c) no research yet tested whether the absence of disease risk alleles mediates/explains the delayed disease onset observed in long-lived families, and d) in silico follow-up analyses that go beyond observing a difference in the number of disease risk alleles are currently scarce. We address these issues by using nationwide causes of death data to identify the most prevalent diseases for which GWASs are constructed. In addition, we use the Longevity Relatives Count (LRC) score to quantify familial longevity, thereby limiting the chance of including phenocopies. Next, we employ time to event models to test if the delayed disease onset observed in descendants of long-lived families is mediated by the absence of disease-associated alleles. Finally, we test which molecular pathways linked to the diseases, also link to prospective survival at later (90+) age, providing valuable insights into healthy aging.

We use data from the Leiden Longevity Study (LLS)^26^ and the Leiden 85-plus study (L85+)^27^. The LLS consists of 651 families covering three generations which we denote filial 1 to 3 (F1-F3), comprising 8,909 persons in total. Inclusion was initiated in 2002 with F2 nonagenarian siblings. Furthermore, the F3 offspring and the respective F3 partners of the offspring were included as well as their family members. The L85+ contains 599 unrelated individuals of 85 years and older. First, we use the nationwide causes of death information provided by Statistics Netherlands (CBS) to identify the diseases responsible for the most deaths in the country. Based on this information, we select the most robust GWASs for those diseases and quantify them into 18 polygenic risk scores (PGSs). Using the LRC score, we investigate whether having an increasing number of exceptionally old ancestors in the F3 generation associates with a lower genetic predisposition to the identified disease PGSs. In addition, we study if the genetic predisposition to these diseases differs between the extremes in the LRC score distribution of the F3 generation. Next, using time to event (survival) models, we test a mediation model to investigate to what extent the genetic predisposition to coronary artery disease (CAD) explains the delayed cardiovascular disease onset in the F3 generation. Finally, to study the molecular pathways underlying the observed CAD - PGS association, we developed a cholesterol pathway-based sub-PGS for CAD which we associate with time to all-cause mortality in the F2 nonagenarian siblings and the L85+.

## Results

### Study Populations

The LLS F3 generation (F3 offspring, N=1671 and F3 partners of the offspring, N=626) were included between 2002 and 2006 at the average age of 59 years. The study inclusion was based on F2 nonagenarian siblings (N=941). Hence, the F3 offspring were included if they had at least one F2 exceptionally old parent and F2 aunt or uncle (females ≥91 years and males ≥89 years). F3 partners were included only if ancestral data about their parents was available. The LLS consists of 651 three-generational families, defined by offspring who have the same parents (mean sibship size is 2.58). From inclusion onward, the F3 generation was followed over time, with a maximum mortality follow-up of 19 years (2002–2021) and maximum morbidity follow-up of 16 years (2002–2018). In 2021, 228 (14%) F3 offspring and 83 (13%) F3 partners were deceased, and 1443 (86%) F3 offspring and 543 (87%) F3 partners were still alive. In 2018, 614 (37%) offspring and 252 (40%) partners had a disease diagnosis whereas 591 (35%) offspring and 216 (35%) partners did not have a disease diagnosis. From inclusion, the nonagenarian siblings were followed over time with a maximum mortality follow-up of 19 years (2002-2021). In 2021, all nonagenarian siblings were deceased, with the oldest person reaching 109.85 years (Figure 1 and Table 1).

**Figure 1.**
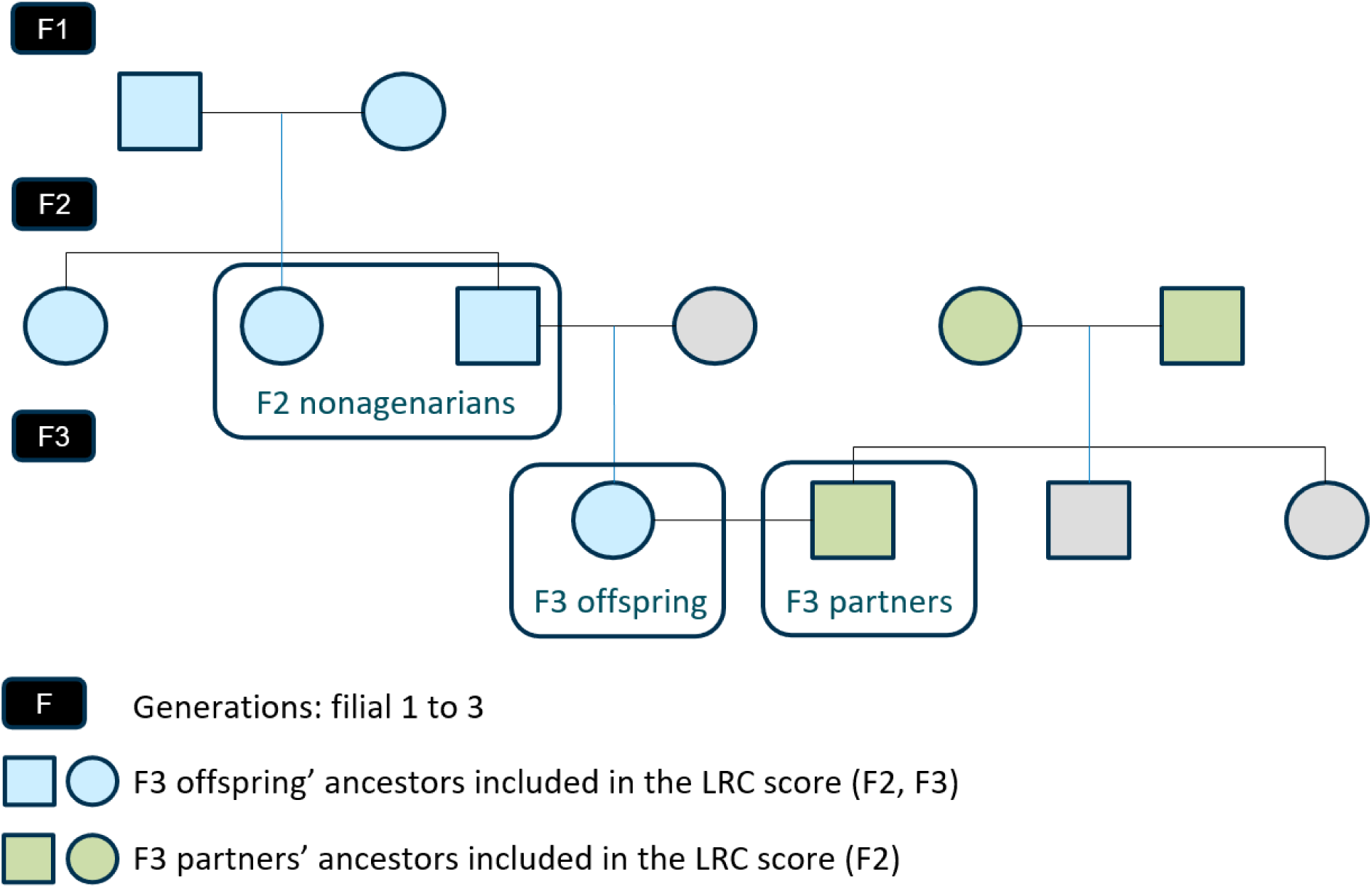
Conceptual pedigree of a 3 filial (F) generation family in LLS. The figure represents a hypothetical family from the LLS covering 3 filial (F) generations. Circles represent women, squares represent men. Groups within a square highlight the individuals considered in the analyses: F3 offspring and F3 partners, and the F2 nonagenarian siblings. Light blue: ancestors of the F3 offspring used to estimate the LRC score. Light green: ancestors of the F3 partners used to estimate the LRC score. Light gray: family members not included in the study.

**Table 1.** Demographic characteristics of the Leiden Longevity Study (LLS) and the Leiden 85-plus study (L85+)

|  | LLS |  |  | L85+ |
| --- | --- | --- | --- | --- |
|  | F3 offspring | F3 partners | F2 nonagenarian siblings | L85+ |
| Number, N (number of families) | 1671 (649) | 626 (626) | 941 (421) | 335 (335) |
| Female, N (%) | 904 (54) | 361 (58) | 591 (63) | 229 (72) |
| Range birth cohorts, years | 1923-1971 | 1925-1974 | 1900 - 1916 | 1912-1914 |
| Alive after follow-up, N (%) | 1443 (86) | 543 (87) | 0 (100) | 0 (0) |
| Deceased during follow-up, N (%) | 228 (14) | 83 (13) | 941 (100) | 334 (99) |
| Missing age, N (%) | 0 (0) | 0 (0) | 0 (0) | 1 (0) |
| Mean ad* or al#, years (SD) | 74.89 (6.65) | 74.74 (7.05) | 93.37 (3.61) | 92.65 (2.91) |
| Genotypes available, N (%) | 1671 (100) | 626 (100) | 941 (100) | 335 (100) |
| Disease status available, N (%) | 1205 (72) | 468 (75) | - | - |
| LRC30%, N (%) | 1389 (83) | 152 (24) | - | - |
| LRC0%, N (%) | - | 474 (76) | - | - |

Leiden Longevity Study (LLS): LLS-offspring and their partners were included in the LLS between 2002 and 2006. For all groups, mortality information was updated in January 2021. All mortality information was obtained from the official and verified Netherlands Population Registers. Disease (morbidity) information was updated in 2018 based input from the General Practitioners of the IPs and their partners. The study covers a maximal mortality follow-up of 19 years (2002-2021) and maximal morbidity follow-up of 16 years (2002-2018). Leiden 85-plus Study (L85+): Participants were included during the month of their 85<sup>th</sup> birthday in the L85+. Mortality information was updated in December 2013. All mortality information was obtained from the civic registry of Leiden. \* ad = age at death and # al = age at last observation.

The participants of the Leiden 85-plus Study (L85+) were included in the month of their 85^th^ birthday^27,13,28^. Participants were followed over time with a maximum mortality follow-up of 12 years (2002-2014). At the end of the follow-up, all participants were deceased, with the oldest person reaching 101.81 years (Table 1). Morbidity follow-up data and ancestral longevity data were not available.

### Increasing number of long-lived ancestors indicates a lower genetic risk for CAD

To investigate whether having an increasing number of exceptionally old ancestors associates with a decreasing genetic risk for the diseases causing most deaths (see Figure S1), we quantified the number of exceptionally old ancestors for the F3 generation using the Longevity Relatives Count (LRC) score^29,30^. We tested 18 PGSs for mortal diseases (statistical model A1 - a) and observed that an increasing LRC score associates with decreasing PGS levels for coronary artery disease (CAD - PGS) (b=-0.047, p=1.0×10^-05^). This result indicates that the more exceptionally old ancestors an individual has, the lower their genetic predisposition to CAD is (Figure 2).

**Figure 2.**
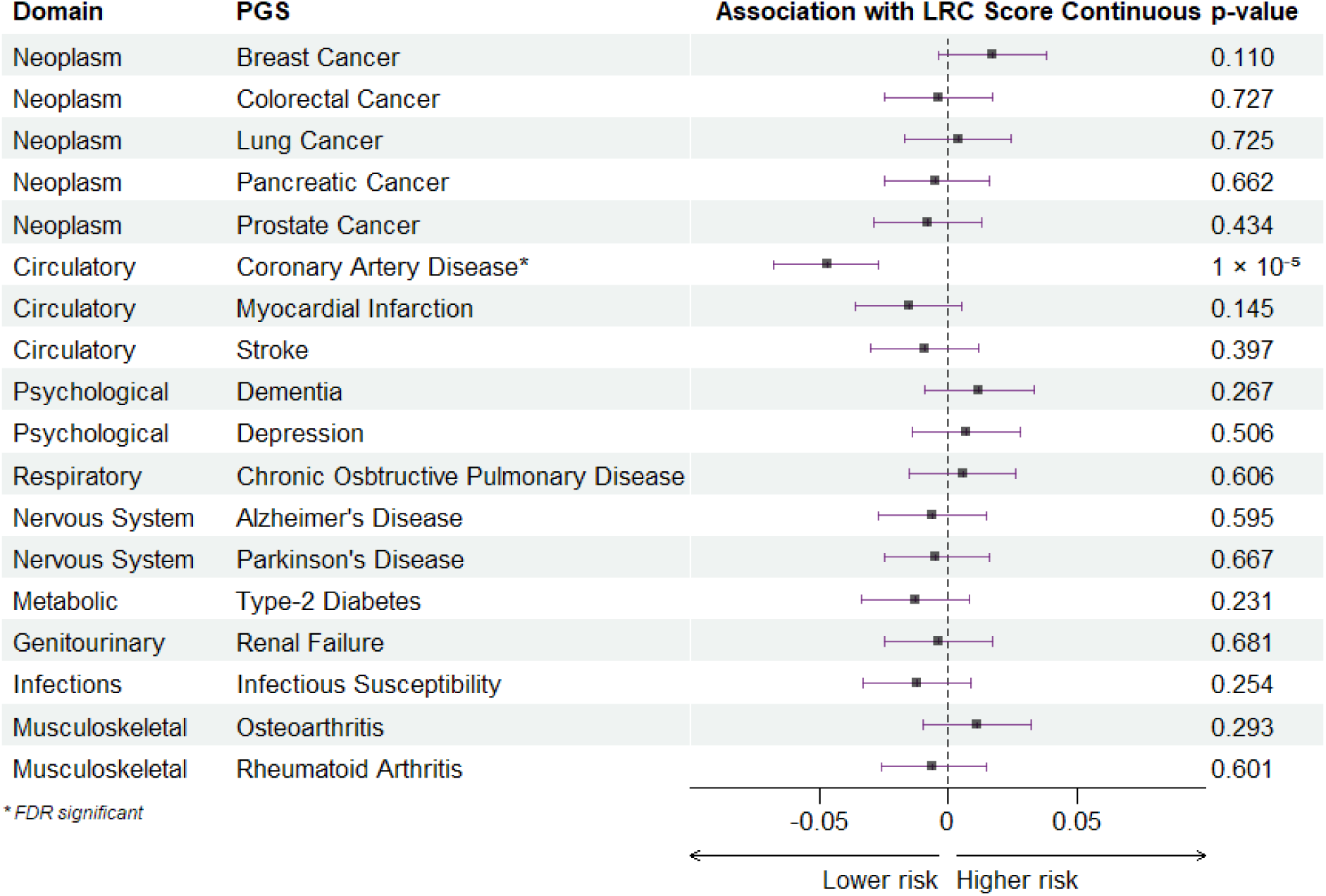
Association between LRC Score Continuous and Polygenic Scores for diseases. The figure shows the association between the Longevity Relatives Count (LRC) Score Continuous and the Polygenic Scores (PGS) for the top 18 diseases responsible for deaths in the Netherlands according to the Statistics Netherlands (CBS). Negative values indicate that for every unit increase in the LRC, the lower the genetic predisposition to the disease is. The asterisk indicates which associations were significant after FDR correction for multiple testing.

To study the extremes of the LRC score distribution as well as non-linear effects, we stratified the F3 generation into two groups: those with 30% or more exceptionally old ancestors (LRC30%) and those without any exceptionally old ancestors (LRC 0%) (statistical model A2 - a). We observed that the LRC30% group had 0.26 (95% confidence interval [CI] 0.37-0.15) standard deviations (SD) lower PGS for CAD than the LRC0% group. For both analyses the effect of CAD remained significant after FDR correction for multiple testing. None of the other PGS differed significantly between the two groups. We found however, the effect of the PGS for myocardial infarction to be borderline significant, pointing in the same direction as CAD - PGS results (Figure S2).

We conducted additional checks to inspect the robustness of the association between the LRC groups (LRC30% and LRC0%) and the CAD - PGS. We estimated the CAD PGS using three different approaches. First, we estimated the CAD - PGS using FDR significant single nucleotide polymorphisms (SNPs) instead of genome-wide significant SNPs (statistical model A2 - b). Second, we estimated the CAD - PGS using the LDPred tool to include non genome-wide SNPs^31^ (statistical model A2 - c). Third, because *APOE* is a gene that has been widely associated with cardiovascular disease as well as healthy aging and longevity^32–34^ (statistical model A2 - d), we recalculated the CAD - PGS without SNPs in or near the *APOE* gene. Using FDR significant SNPs, the association between the LRC groups and CAD - PGS is also significant (b= - 0.22, CI - 0.33 - - 0.11), but slightly smaller than when using genome-wide significant SNPs. LDPred-based results indicated no difference between the two groups (b=0.01, CI - 0.09 - 0.07), suggesting that the CAD most significantly associated SNPs contribute to the difference between the LRC30% and LRC0% groups. Finally, when removing SNPs near the *APOE* gene, the effect was slightly smaller but of similar magnitude (b=- 0.24, CI - 0.35 - - 0.13), showing that a significant portion of the difference in CAD – PGS between LRC30% and LRC0% is driven by SNPs near other genes than *APOE* (Table S1).

### Delayed CVD onset in familial longevity is mediated by a lower genetic risk for those diseases

Next, we investigated whether the lower genetic predisposition to CAD, observed in the LRC30%, mediates cardiovascular disease (CVD) incidence. As the SNPs identified in the CAD - GWAS^35^ underly broader cardiovascular health, our analyses were set out in three steps. In the first model (statistical model B1 - a; Figure 3 M1), we investigated to what extent the CAD - PGS associates with prospective CVD onset. Then, in the second model (statistical model B1 - b; Figure 3 M2), we investigated if the LRC30% and LRC0% groups differ in time to CVD onset. Finally, we investigated if the difference in CVD onset between LRC30% and LRC0% could be explained by the CAD - PGS (statistical model B1 - c; Figure 3 M3). To answer these questions, we conducted Cox frailty (random effect) regression analysis (statistical models B1) and validated the results using Accelerated failure time (AFT) models (statistical models B2).

**Figure 3.**
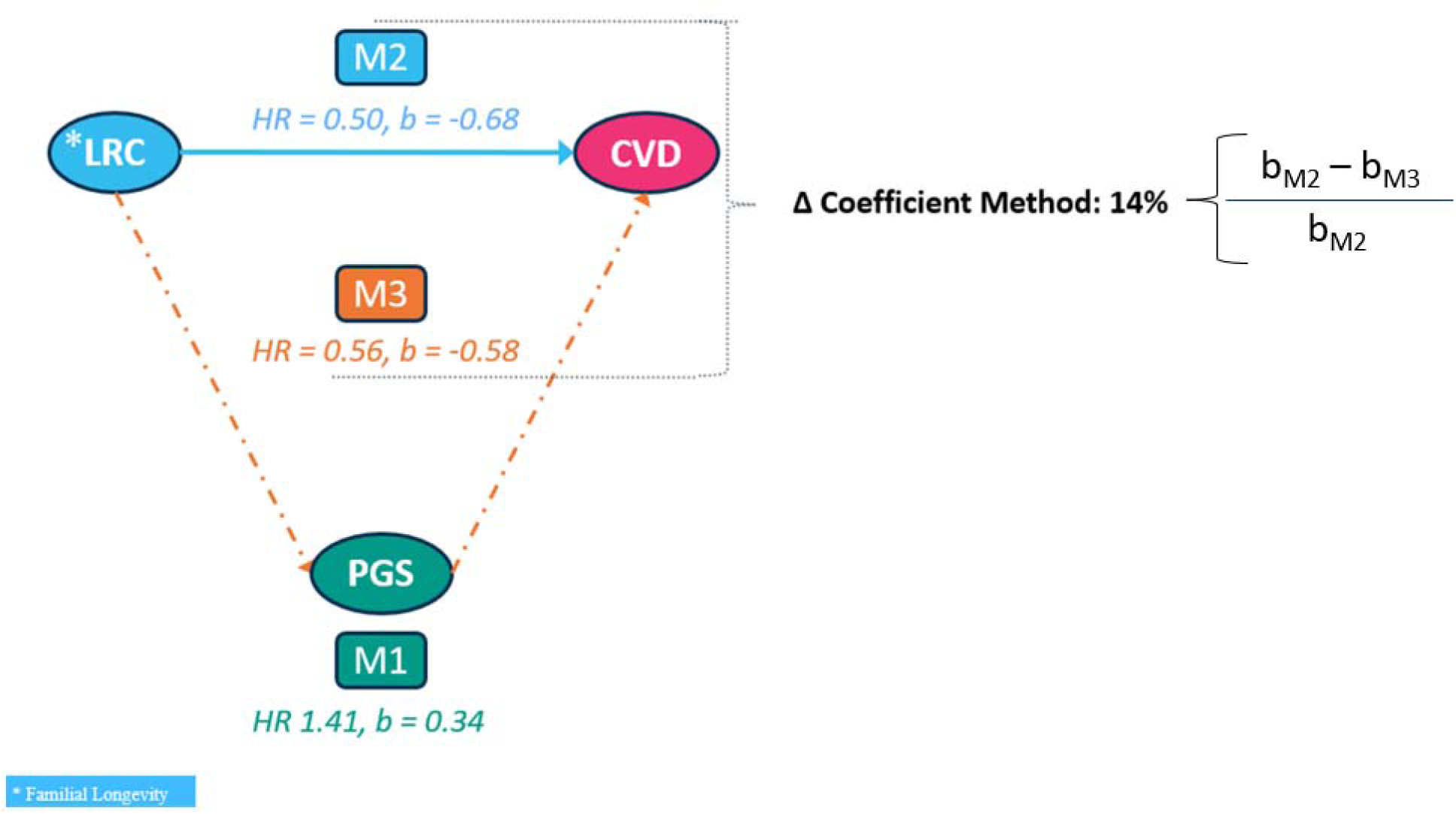
Genetic predisposition to CAD explains delayed CVD incidence. The figure shows the association between LRC score groups and cardiovascular disease (CVD) incidence in model 1 (M1), the association between the genetic predisposition to coronary artery disease (CAD) captured by the CAD polygenic score and CVD incidence in model 2 (M2), and the association between LRC score and CVD mediated by the genetic predisposition to CAD in model 3 (M3). The mediating effect of the genetic predisposition to CAD is quantified using the difference of coefficients method (ΔCoefficient Method).

In the first step, results point to an association between genetic propensity to CAD and CVD incidence. Every standard deviation (SD) increase in the CAD - PGS was associated with a 41% (b=0.34, CI=1.15-1.72)) increase in CVD incidence. We further observed that individuals in the lower tertile for the CAD - PGS had a delayed onset of CVD of almost 10 years in comparison to individuals in higher tertiles (Figure S3). In the second step, we observed that the LRC30% group had a 50% (CI=0.33-0.78) reduced yearly onset of CVD compared to the LRC0% group. In the third step, the mediation model was fitted using the delta coefficient method^36^. We compared the beta from the second model (b=- 0.68) and from the third model (b=- 0.58) and observed that 14% (0.10/0.68=0.14) of the delayed CVD incidence in the LRC30% group was explained by their lower genetic predisposition (less disease risk alleles) for CAD (Table S2).

We triangulated our results to ensure that our findings are robust for the non-collapsibility property of Cox-type models, which can affect mediation analysis. Hence, we fitted a parametric AFT model which is robust for this property (see methods)^37^. We observed a Time Ratio (TR) of 0.81 (CI=0.72-0.92), indicating that with every SD increase in CAD - PGS, the cardiovascular disease incidence is accelerated by 19%. Note that higher HRs and lower TRs indicate an increased risk. In addition, the incidence of cardiovascular disease is decelerated by 46% (TR=1.54, CI=1.15-2.07) for the LRC30% group compared to the LRC0% group. Lastly, the AFT-based mediation analysis pointed to a mediation effect of 20% (0.09/0.43=0.20), slightly stronger than observed in our Frailty-Cox analyses. The results in our triangulation are in concordance, indicating that the delayed onset of CVD observed in the LRC30% group when compared to the LRC0% group can be partly explained by the lower genetic predisposition to CAD observed in the LRC30% group (Figure S4 and Table S3).

### CAD-based gene-annotation enrichment analysis indicates potential longevity pathways

To better understand the biological pathways by which the CAD - PGS is involved in longevity, we conducted a gene-annotation enrichment analysis of the genes composing the CAD - PGS using the Database for Annotation, Visualization, and Integrated Discovery (DAVID)^38,39^. As input, we provided the 207 unique genes which mapped to the 250 SNPs of the CAD - PGS (Table S4). We identified seven clusters with an enrichment score of at least 3 that mapped to 99 genes and 132 SNPs respectively. The seven clusters were: a) focal adhesion, b) Age-RAGE, c) RAP-RAS-MAPK, d) Rho GTPase, e) Cholesterol metabolism, f) Platelet degranulation, and 7) Epidermal Growth Factor signaling (Figure S5).

### Cholesterol metabolism pathway-based-PGS associates with mortality in later age

Because longevity GWAS yielded only a handful of consistent findings^44–46^, we examined whether the pathways from our enrichment analysis are associated with later-age (90+) prospective survival. To this mean, we constructed three “pathway-PGSs” based on the 132 SNPs from the enrichment analysis. In the first pathway-PGS we incorporated all the 132 SNPs into a All - Clusters - PGS (Table S5). Next, since research has shown that cholesterol metabolism plays an important role in aging and mortality^40–43^, we constructed the second pathway-PGS representing the Cholesterol Metabolism cluster (ChoMet_52_ - PGS) based on 30 genes (out of the mapped 99 genes) and 52 SNPs (out of the 132 SNPs; Table S6). Finally, we used the remaining 80 SNPs (mapping to the other 69 genes), to construct the third pathway-PGS representing the other 6 clusters (6 - Clusters - PGS; Table S7).

Analyses were performed in the nonagenarian siblings and replicated in the L85+. Given the sibship relations present in the LLS data, we used random effect (frailty) cox-type regression model (statistical models C1) to investigate the association between each pathway-PGS and time to mortality. For the replication using the L85+ data, we used a regular cox-type regression model (statistical models C2).

In the F2 nonagenarian siblings, we observed that the yearly risk of dying increases by 8% (HR=1.08, CI=1.00-1.16) for every standard deviation increase in the All - Clusters - PGS. This indicates that at 90+, carrying fewer CAD related disease risk alleles is associated with improved survival chances. For the ChoMet_52_ - PGS, we observed that the yearly risk of dying increased by 12% (HR=1.12, CI=1.04-1.20) for every standard deviation increase. This translates to a difference of 2.5 years in the median survival between the individuals in the lower tertiles (lower tertile plus middle tertile) compared to the higher tertile. The PGS based on the remaining 6 clusters, showed non-significant effect (HR=0.98, CI=0.91-1.05). In the L85+ participants, we observed similar results for the All - Clusters - PGS and 6 - Clusters - PGS but with a slightly attenuated effect. For the ChoMet_52_ - PGS we observed similar results: the yearly risk of dying increased by 11% (HR=1.11, CI=1.00-1.23) for every standard deviation increase. The difference in the median survival between individuals in the lower tertiles and higher tertile is almost one year (Figure 4, Table S8). Finally, we conducted a meta-analysis with the results observed for the ChoMet_52_ - PGS in the F2 nonagenarian siblings and L85+ participants. Using a pooled common effects (fixed effects) model we observed that for every SD increase in the ChoMet_52_ - PGS, the yearly chance of dying increased by 12% (HR=1.12, CI=1.05-1.18, p=0.0003; See Figure S6).

**Figure 4.**
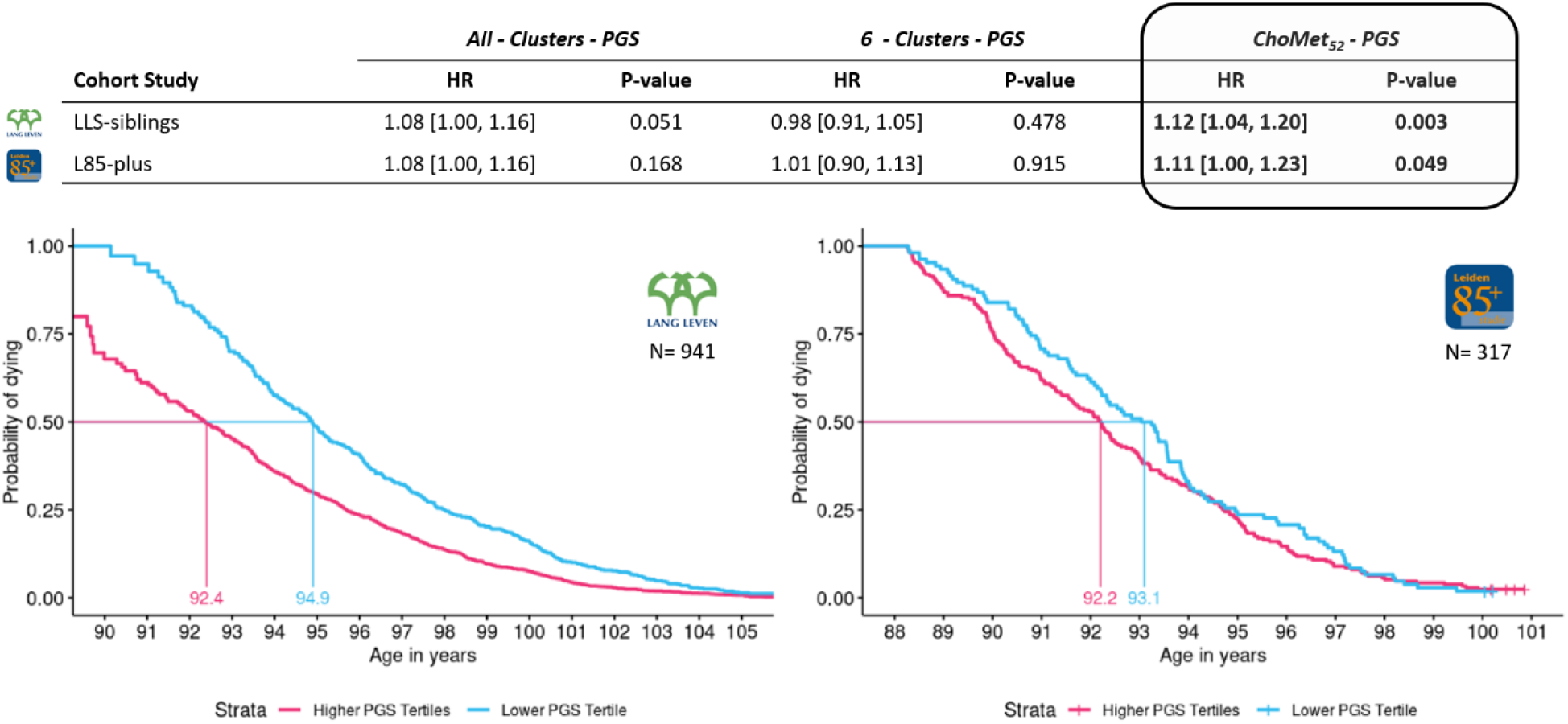
Cholesterol Metabolism PGS associates with Mortality. The figure shows the association between the pathway polygenic score for cholesterol metabolism (ChoMet_52_-PGS) and mortality in the Leiden Longevity Study (A) and in the Leiden 85-plus Study (B). Participants in the higher tertiles for the ChoMet_52_-PGS have are more likely to dye 2,5 years earlier in the Leiden Longevity Study and almost a year earlier in the Leiden 85-plus Study.

We also performed a robustness check, removing SNPs with a likely functional impact on the *APOE* gene due to its strong association with the onset of cardiovascular disease, healthy aging, and longevity^32–34^. Out of the 52 SNPs present in the ChoMet_52_ - PGS, 3 act on the *APOE* gene (rs7421, rs429358, and rs8108474). We therefore constructed a revised ChoMet_49_ - PGS with the remaining 49 SNPs and verified if the association with mortality still held. In the F2 nonagenarian siblings, the yearly risk of dying increased by 9% (HR=1.09, 95% CI=1.01-1.18) for every standard deviation increase in the revised ChoMet_49_ - PGS (ChoMet_49_ - PGS), while this was 5% (HR=1.05, 95% CI=0.95-1.17) in the L85+ participants. Meta-analysis on the results indicated that for every SD increase in the ChoMet_49_ - PGS, the yearly risk of dying increased by 8% (HR=1.08, 95% CI=1.01-1.14, p=0.019). We observed a reduction in the pooled effect of the meta-analysis from 12% (ChoMet_52_ – PGS) to 8% (ChoMet_49_ - PGS). Though the effect is slightly mitigated, this indicates that the association with mortality still holds beyond the contribution of *APOE*-related SNPs (Figure S7).

## Discussion

In the present study, we have shown supporting evidence for a lower genetic predisposition to CAD for descendants of exceptionally long-lived families. Focusing on these persons, our results indicated that the more exceptionally old ancestors the descendants had, the lower their genetic predisposition to CAD. Moreover, the genetic predisposition to CAD explained between 14% to 20% of the delayed cardiovascular disease onset observed in the descendants of exceptionally long-lived families. Finally, we showed that lower genetic risk for cardiovascular disease, particularly through the cholesterol metabolism pathway, represents a key biological mechanism contributing to survival at a later age. Using two independent samples (LLS and L85+), we showed that, at age 90+, for every SD increase in our pathway cholesterol metabolism polygenic score, the yearly risk of dying increased by 12%, indicating that having fewer CAD disease risk alleles are associated with improved survival chances. This amounts to a difference of 2.5 years in median survival between those in the lowest genetic risk tertile compared to the highest tertile. Our results highlight that in healthy aging the absence of CAD risk alleles and cholesterol metabolism is a key-feature. Furthermore, we showed how incorporating the familial component of longevity can further enhance the study of chronic diseases and healthy aging.

Our multigenerational approach focusing on intergenerational transmission of healthy survival up to very high ages showed, for the first time, the additive association between having an increasing number of exceptionally old ancestors and the genetic predisposition to CAD. Moreover, this is the first study that follows-up on these results by 1) testing whether the absence of genetic disease risk explains the delayed disease onset observed in descendants of long-lived families, 2) conducting a gene-annotation enrichment analysis to construct mechanism specific PGSs, and 3) link these mechanism specific PGSs to survival above age 90 in two independent datasets. Previous research also pointed to the link between CAD and longevity. These studies observed an inverse association between genetic predisposition to CAD and longevity in exceptionally old individuals^23–25^ as well as to being an offspring of a longer-lived parent^47^. Contrary to previous research, we did not observe an association with genetic predisposition to any other diseases. We note, however, important differences. First, we focused on offspring of increasingly long-lived ancestors while most studies^23–25^ compared exceptionally old individuals to younger controls, many of whom were not from the same generation and without taking family predisposition into account. Our results more directly line-up with Pilling et al.^47^ who focused on the offspring of long-lived parents using data from UK-Biobank. Beyond CAD, they also found an effect for other cardiovascular related traits as systolic blood pressure. However, they focus on parental age, not extended ancestral mortality, which was reported by the study participants instead of their parents^29^. Another difference concerns the selection of diseases and traits. We based our selection on disease domains with the highest mortality burden to the population while previous studies included either a few diseases^23^ or a combination of diseases^24,25,47^, behavioral traits^24,25^, and metabolic indexes^24,47^.

This study focuses on the absence of disease-risk alleles as a key feature of reaching very old age in good health. We showed that a greater number of long-lived ancestors is additively associated with lower genetic risk for CAD, and our mechanistic analyses demonstrated that this reduced genetic predisposition to CAD accounts for 14–20% of the delayed onset of cardiovascular disease in the descendants of exceptionally old persons. These results come with a few implications. First, the CAD - PGS offers possibilities to dive into alleles more closely linked to healthy aging but understanding of how related pathways act remains a topic for subsequent research. Second, there is still a large portion of the difference in CVD disease onset between the descendants of exceptionally old individuals and their partners with no exceptionally old ancestors that could be explained either by other disease-related alleles not captured in this study, rare and structural variants^48^, including protective alleles as recently observed in the LLS^49^, and/or by socioeconomic and behavioral indicators^9,50^. Given that the SNPs identified in the CAD - GWAS underly general cardiovascular health, a strong link to the onset of CVD would be likely. The strong association between the CAD – PGS and the onset of CVD observed in our study corroborates the expectation. Inclusion of additional specific CVD related SNPs to the CAD – PGS could potentially increase the association with CVD onset or the construction of a CVD – PGS. In addition, descendants of exceptional long-lived persons may benefit from healthier environments provided by their parents as posited by health selection theory^51^. Healthier individuals are more likely to achieve higher socio-economic status because early health advantages enable better educational attainment, occupation, and wealth. Exceptional old parents may provide prolonged support to their descendants through the life course. Studying the unique environment and the gene-environment interplay are the next steps in understanding the link between longevity and healthy aging.

Lower CAD genetic risk was linked to healthy aging, and a cholesterol metabolism PGS (ChoMet-PGS) predicted time to all-cause mortality in two long-lived cohorts, even after excluding *APOE*-related SNPs. This result is unique since it is hypothesized that the cholesterol metabolism pathway is linked to human longevity mainly via *APOE*, a gene identified in many genetic longevity studies. Owning to no model organism-based genetic study having implicated cholesterol metabolism in the regulation of longevity, the link is mainly inferred from the role the different *APOE* isoforms play in humans^19,52^. Our results suggest that other SNPs involved in cholesterol metabolism are linked to survival at a later age, potentially acting on genes connected to lipoprotein particle binding. Having a lipoprotein signature characterized by the presence of higher LDL particles and lower triglycerides’ levels was previously associated with longer lifespan in previous studies with the LLS^53,54^. Cholesterol and lipid metabolism could be new avenues to explore in unrevealing molecular functions linked to healthy aging.

Our study design, using multigeneration familial data, provides a unique combination of nationwide registry-based mortality data, disease information obtained from the general practitioners of the participants, and genomic data. This unique combination made it possible to employ in silico follow-up analyses, offering a mechanistic perspective on the implications that having fewer disease risk alleles have on exceptionally old individuals or its family members. Our results may encourage future genetic studies to incorporate ancestral data to genomic data to foster new discoveries. Despite the robustness of using multigeneration familial data, our study comes with limitations. We used the third-generation partners to construct our comparison group with no exceptionally old ancestors. Although our design controls for shared environmental effects, it cannot completely do so. Since our comparison groups share the same household, we cannot rule out potential underestimation of the difference between the descendants and their partners. Hence it would be still necessary to replicate our analysis with general population unrelated individuals as the comparison group. In addition, our study was the first to employ the LRC score, thus replications with other cohort studies from other exceptionally old individuals from other ancestries with access to ancestral mortality data is necessary. There may be several reasons why we did not pick up the effect of other PGSs than the CAD - PGS. One reason concerns the power of some of the most recent GWASs which may have impacted the estimate of the genetic predisposition to disease via PGS. Also, the use of penalized regression methods including non-significant SNPs may introduce noise as observed in our study and Hu et al.^24^. In addition, using data from larger multigenerational cohort studies than the LLS may detect smaller effects. Finally, our results are based on European ancestry. Before any generalizations can be extended to other populations, replications in other ancestry samples must be conducted.

In conclusion, our novel family-based study design shows that descendants of exceptionally long-lived families have a lower genetic predisposition to CAD. The association was independent of the presence of *APOE* related SNPs. When removing SNPs near the *APOE* gene, we observed a lower genetic predisposition to CAD of similar magnitude, suggesting that there are other CAD related genes associated with familial longevity. A lower genetic predisposition to CAD partially explains the later onset of CVD for those individuals, highlighting that the absence of CAD related alleles seem relevant for healthy aging. The non-explained portion hints to residual genetic effects, environmental effects, and gene-environment interactions. Future study efforts should focus on studying the influence of environmental factors on those who have exceptionally long-lived ancestors and the ones with no exceptionally long-lived ancestors. Given that the individuals who reach older ages in good health tend to be the ones with higher socioeconomic status^50^, examining a potential casual link between health and socioeconomic opportunities emerges as topic for future research. Finally, we have shown that the cholesterol metabolism pathway has a central role in healthy aging from midlife to later age, pointing it as a key topic for investigation in future studies. Further investigations on this pathway may lead us to a better understanding on how to make healthy aging a reality for all.

## Methods

### Leiden Longevity Study (LLS)

The LLS consists of 651 three-generation families (F1-F3) and was initiated in 2002 to identify mechanisms involved in healthspan and exceptional survival. Inclusion took place between 2002 and 2006, starting with the recruitment of living sibling pairs (F2 nonagenarian siblings), which compose the LLS second generation (F2). The age criterion for inclusion within the F2 nonagenarian siblings’ pairs was 89 years or older for males and 91 years or older for females. Furthermore, the parents of the F2 nonagenarians had to be of Dutch Caucasian ancestry for inclusion. Subsequently, the LLS was extended to the third generation (F3), with the inclusion of the F3 offspring of the nonagenarian siblings and the F2 partners of the offspring. The F3 offspring were included if they had at least one exceptionally old parent and aunt or uncle (males ≥ 89 years and females ≥ 91 years). From inclusion onward, the F3 offspring were followed over time, with a maximum mortality follow-up of 19 years (2002–2021) and maximum morbidity follow-up of 16 years (2002–2018). In 2018, 186 (11%) F3 offspring and 67 (11%) of F3 partners were deceased, and 1485 (89%) of F3 offspring and 559 (89%) F3 partners were still alive with 1205 (72%) of the F3 offspring and 468 (75%) of the F3 partners with a disease diagnosis available. In 2021, 228 (14%) F3 offspring and 83 (13%) partners were deceased, and 1443 (86%) F3 offspring and 543 (87%) F3 partners were still alive.

In the present study, we first focus on the F3 generation. To map the familial history of longevity, data from the Ancestral Groups of the F3 generation was used. For the F3 offspring it was used data from the F1 grandparents (N=760), the F2 parents of the offspring (N=1295), and the F2 aunts or uncles (N=2364). For the F3 partners it was used data from the F2 parents of the partners (N=1252), as input for the LRC score (See Table S9)

In the last section, the analysis is then focused on the F2 nonagenarian siblings (either a mother or father from F3 offspring and respective sibling) which were identified in the present study if genotyped data was available (Figure 1 and Table 1).

### Leiden 85-plus Study (L85+)

The L85+ started in 1986 with gerontologic surveys of the population of the oldest living individuals of the municipality of Leiden, The Netherlands. Between September 1997 and September 1999, members of the 1912 to 1914 birth cohort (705) were eligible to participate in the study but only 599 individuals were enrolled in the month of their 85th birthday with follow-up (See der Wiel et al.^27^, Postmus et al.^28^). Of these 599 individuals, we used demographic and genotyped data available at the time, a subset of 335 individuals who reached the age of 88 years which comprised the replication sample for the association between mortality and pathway polygenic scores (Table 1).

### Genotyping

In the LLS, genotyping was performed at baseline as described in detail in Beekman et al.^55^ with the Illumina Human660W and Illumina OmniExpress arrays (Illumina, San Diego, CA, USA). Genotype imputation was performed using 296,486 SNPS with SNP-wise call rate (>95%), minor allele frequency (>1%) and no derivation from the Hardy–Weinberg equilibrium (p-value > 1×10^-4^) at the Michigan Imputation Server^56^ with Haplotype Reference Consortium (HRC) reference panels (HRC1.1). DNA genotyping for L85+ was performed at baseline as described in Deelen et al. and Postmus et al^28,57^ with Illumina Human660W-Quad and OmniExpress Bead Chips (Illumina, San Diego, CA, USA). Genotype imputation was performed in the Michigan Imputation Server 2^56^ with Haplotyped Reference Consortium (HRC) reference panels (HRC-r1.1, hg19). Imputation was performed using 601,010 SNPs with SNP-wise call rate (>95%), minor allele frequency (>1%) and no derivation from the Hardy–Weinberg equilibrium (p-value > 1×10^-4^). For an overview see Table S10 and Table S11 in the Supplementary Materials.

### Mortality information

In the LLS, mortality information was verified by birth or marriage certificates and passports whenever possible. Additionally, verification took place via personal cards which were obtained from the Dutch Central Bureau of Genealogy (*Centraal Bureau voor Genealogie*, CBG). In January 2021, all mortality information was updated through the Personal Records Database (*Basisregistratie Personen*, PRD) which is managed by Dutch governmental service for identity information (https://www.government.nl/topics/personal-data/personal-records-database-brp). In the L85+, mortality information was verified in the civil registry of the city of Leiden. By February 2014, all the participants were deceased.

### Disease information

Disease data for the LLS has been retrieved from the General Practitioners (GPs) of the F3 generation and covers the period from birth until 2018. GPs extracted the presence of chronic age-related diseases and the year the disease occurred from their electronic health records. The GP records are kept up to date when a person switches from one GP practice to another. Based on the records, we constructed the CVD outcome group based on the International Classification of Diseases, 10^th^ revision (ICD-10) of Transient Ischemic Attack (TIA, I60-I69), Cerebrovascular Accident (CVA, I63), Angina Pectoris (AP, I20), and Acute Myocardial Infarction (MI, I21).

### Lifetables

In the Netherlands, population-based cohort lifetables are available from 1850 until 2021^58^. These lifetables contain, for each birth year and sex, an estimate of the hazard of dying between ages x and x + n (hx) based on yearly intervals (n=1) up to 99 years of age. Conditional cumulative hazards (Hx) and survival probabilities (Sx) can be derived using these hazards. In turn, we can determine to which sex and birth year survival percentile each person of our study belonged to. For example: a person was born in 1876, was a female, and died at age 92. According to the lifetable information this person belonged to the top three percent survivors of her birth cohort, meaning that only three percent of the women born in 1876 reached a higher age. We used the lifetables to calculate the birth cohort and sex specific survival percentiles for all persons in the LLS. This approach takes sex-specific differences in longevity into account and prevents against the effects of secular mortality trends over the last centuries and enables comparisons across study populations^17,59^. In the LLS some ancestors (only aunts/uncles) were still alive (right censoring). To deal with non-extinct birth cohorts, we used the prognostic lifetables provided by Statistics Netherlands^58,60^ and to deal with right censoring we used single imputation where we estimated an age of death based on the remaining life expectancy at the age of censoring.

### Longevity Relatives Count Score

The Longevity Relatives Count (LRC) score was used in the LLS to map the family history of longevity in the F3 generation. The LRC score indicates the proportion of ancestors that became long-lived, weighted by the genetic distance between an individual and their ancestors. For example, an LRC of 0.5 indicates 50% long-lived ancestors. For this study, two generations of ancestors were available to calculate the LRC score for the F3 offspring and one generation for the F3 partners (See Figure 1). To estimate the LRC score, mortality information was used as updated in 2021. A detailed description of the score and respective methodology employed is depicted in van den Berg et al.^30^ and can be calculated as follows:

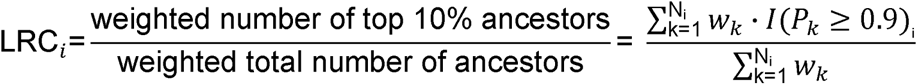

Where *i* refers to the person for whom the score is built. k is an index referring to each ancestral blood relative (from here: ancestors) of person i who are used to construct the score. Ni refers to the total number of ancestors of person *i*, P_k_is the sex and birth year-specific survival percentile, based on lifetables of ancestor k, and l (P_k_≥ 0.9) indicates if ancestor k belongs to the top 10% survivors. 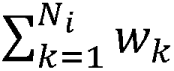 is the weighted total number of ancestors of person *i*. The relationship coefficients are used as weights w_k_.

### Polygenic Scores

#### Causes of death

Using aggregated data from Statistics Netherlands (CBS), we identified all causes of death domains based on diseases using the ICD-10 overall groupings. Next, we excluded the domains that were specific to a time-period or not relevant for genetic analyses: a) COVID-19 (Coronavirus disease 19), b) external causes of morbidity and mortality, c) unclassified symptoms, signs, and abnormal findings. Finally, we identified the top 10 domains based on the proportion of mortality within a domain compared to the total mortality for the years 2021, 2022, and 2023 (Figure S1). The CBS data shows that on average, 170k individuals passed away in the Netherlands from 2021 to 2023. The top 10 causes of death domains covered respectively 77%, 82%, and 84% of deaths in this period. To go from causes of death domains to specific diseases causing most deaths, we identified the top 5 specific diseases causing most deaths for each domain (see Figure S8 and Table S12 in the Supplementary Materials).

#### Polygenic Risk Score calculation

We matched each specific identified disease responsible for the most deaths to a representative GWAS. The studies were prioritized based on a) the largest number of independent, genome-wide significant SNPs (*p* = 5 x 10^-8^), b) the largest sample size, and c) sufficient information on independent (as derived using LD-based clumping), genome-wide significant SNPs. In particular, the studies needed to report the rsID, chromosome, position, effect/other allele, effect size (beta/odds-ratio), and p-value. If this information was not available, we omitted the study and returned to the initial step (a). In total, 18 GWASs were matched, representing 9 causes of death domains and 27 specific causes of death^35,61–77^. Note that the total number of GWAS does not match the number of specific causes of death. This is because some specific causes of death could not be identified or were captured by a single GWAS. For example, cerebral infarction (IDC10-I36) and Intracerebral hemorrhage were captured as “stroke” (See an overview in Table S12)

In addition to pre-imputation curation of the genotype data as mentioned in the genotyping paragraph, we removed SNPs with an imputation quality lower than 0.3^78,79^. Specifically for the Leiden 85-plus sample (a sample of unrelated individuals), we removed individuals with PIHAT ≥ 0.125 (as depicted in Table S11). Table S13 in the Supplementary Materials shows each disease-PGS and the respective number of SNPs before and after QC. PGS were calculated as follows:

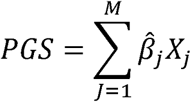

Where the PGS for a single specific cause of death is obtained as the sum of the effect size 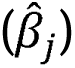 for every SNPs multiplied by its dosage (x_j_). Dosages are coded to represent the risk increasing allele, meaning that a PRS of 0 indicates that a study participant does not have any risk increasing alleles.

#### Pathway Polygenic Scores Construction

To investigate potential mechanisms embedded in the CAD - PGS, we constructed pathway PGS based on pathway clusters identified with the Database for Annotation, Visualization, and Integrated Discovery (DAVID)^38,39^. First, we identified the unique genes with the highest association rank (207 genes) based on the 250 SNPs from the CAD - GWAS by Aragam et al.^35^ (for details, see Supplementary Table 25 in Aragam et al.^35^). Next, the 207 genes were curated (e.g. gene names were updated) and uploaded into DAVID, where the three most appropriate pathway databases were selected (KEGG, Reactome, and WikiPathways) using the *Functional Annotation Clustering* option. We identified seven clusters (Focal Adhesion, Age-RAGE, RAP-RAS-MAPK, Rho GTPase, Cholesterol Metabolism, Platelet Degranulation, Epidermal Growth Factor Signaling) with an enrichment score ≥3. These seven clusters mapped to 99 genes and 132 SNPs which formed the basis for our three novel polygenic scores. The first incorporated all the 132 SNPs into a seven clusters PGS (All - Clusters - PGS). The second PGS was based only on the 52 SNPs (30 genes) of the cholesterol metabolism cluster (ChoMet_52_ - PGS). Finally, we incorporated the remaining 80 SNPs (69 genes) into a third PGS (6 - Clusters - PGS). An overview of the procedure is provided in Figure S5, and the specific SNPs included in each pathway - PGS is available in tables S6, S7, and S8 in the in the supplementary materials.

## Statistical analyses

Statistical analyses were conducted using R version 4.2.2^80^. We reported 95% confidence intervals (CIs) and considered two-sided p-values statistically significant at the 5% level (α=0.05). A list of used R-packages and version numbers will be made available on GitLab (see code availability statement). Linear, Cox-type, Accelerated Failure Time random effects models were used to adjust for within-family relations, assuming family-specific random effects. Families were defined as the F3 offspring or F2 nonagenarian siblings who share the same parents. The replication performed in the L85+ sample was based on unrelated individuals; thus, no random term was included in the model.

### Association between LRC score and genetic predisposition to diseases

*Analysis 1:* To investigate whether an increased number of exceptionally old ancestors associate with having a lower predisposition to the diseases that cause most deaths quantified by PGS, we used mixed-model linear regression analysis (statistical model A1):

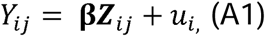

where Y_ij_ is the response (each of the 18 PGS representing the diseases that cause most deaths) person *j* in family *i*. β is a vector of regression coefficients for the main effects of interest (z) which corresponds to the familial longevity captured by the LRC score (which is multiplied by ten for the analyses). The unobserved family-specific random effects u_i_ were assumed to follow a normal distribution (statistical model A1 – a).

To investigate whether there is a difference between the extremes of the LRC groups in genetic predisposition to diseases that cause most deaths quantified by PGS, we used mixed-model linear regression analysis (statistical model A2):

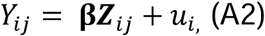

where Y_ij_ is the response (each of the 18 PGS representing the diseases that cause most deaths) person *j* in family *i*. β is a vector of regression coefficients for the main effects of interest (z) which corresponds to the familial longevity captured by the LRC score (which is binarized for the analyses). The unobserved family-specific random effects u_i_ were assumed to follow a normal distribution (statistical model A2 – a).

Robustness checks for the CAD - PGS were conducted. We a) calculate the CAD - PGS based on LDPreD^31^(statistical model A2 - b), b) calculate the PGS using FDR significant SNPs instead of using the genome-wide significant SNPs (statistical model A2 - c), and c) calculate the CAD - PGS without SNPs in or near the *APOE* gene (*APOE* as nearest gene) (statistical model A2 - d). Applying the same equation (2), now β is a vector of regression coefficients for the main effects of interest (Z) which corresponds to the LDPreD-based CAD - PGS (A2 - b), the FDR significant SNPs-based CAD - PGS (A2 - c), and the CAD - PGS without *APOE*-associated SNPs (A2 - d. Other terms of the expression remained the same as noted in 1.

### Associations with CVD incidence

*Analysis 2*: We investigated whether the CAD - PGS associates with cardiovascular disease (CVD) incidence. The CVD outcome group is constructed as a composite based on the ICD-10 coding of TIA (I60-I69), CVA (I63), AP (I20), and MI (I21). Next, we tested if there is a difference in CVD incidence between the LRC groups. Finally, we investigated to what extent the association between the LRC groups and delayed CVD incidence is explained by the CAD - PGS. We used random effect (frailty) Cox regression analysis (statistical model B1):

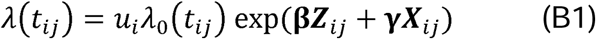

where t_ij_ is the age at cardiovascular disease for person *j* in family *i*. λ_O_ct_ij_) refers to the baseline hazard, which is left unspecified. β is a vector of regression coefficients for the main effects of interest, representing the fixed effect of the variable z which correspond to a) CAD – PGS (B1 – a) and b) familial longevity as grouped using the LRC score (the score was binarized for this analysis, B1 - b). The models are adjusted for left truncation given by the age at entry in the study and confounders (sex, number of diseases pre-inclusion, and medication use at inclusion) as denoted by x in expression 2. u_i_ > 0 refers to an unobserved random effect (frailty) shared by the members of the same family *i* and was assumed to follow a gamma distribution. The mediating effect of CAD - PGS for the association between LRC groups and delayed CVD incidence was quantified using the difference of coefficients method (B1 – c)^36^. The effect is quantified by subtracting the coefficient of the mediation model (B1 - c: b_M3_) from the coefficient of the association between the LRC groups and CVD incidence (B1 - b: b_M2_) and then dividing by the coefficient of the association between the LRC groups and CVD incidence (B1 - b: b_M2_). See overview in Figure 3.

Robustness checks were conducted by replicating analysis 2 with the aim of confirm the mediating effect of CAD - related risk alleles. We used Accelerated Failure Time (AFT) survival analysis, which is a parametric survival model, directly modeling the outcome (time to CVD) instead of operating in the hazard scale. Using the Akaike Information Criterion (AIC) and Bayesian Information Criterion (BIC), we determined the loglogistic distribution as the best fitting distribution of the baseline hazard (See Table S14). The following equation represents the AFT model (statistical model B2):

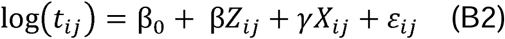

where t_ij_ is the time to event (or survival time, e.g. age at cardiovascular disease) for person *j* in family *i*. β_0_ is the intercept (log-scale baseline time to event). z_ij_ refers to the main covariates of interest for person *j* and family *i* which correspond to a) CAD – PGS (B2 – a) and b) familial longevity LRC score (B2 - b). x_ij_ corresponds to the confounders included in the model (sex, number of diseases, and medication). β, γ are the regression coefficient vectors. ε_ij_ is the random error with a specified distribution (e.g., extreme value for Weibull, logistic, log-logistic, etc.). The mediating effect was quantified by the difference of coefficients method as depicted before (B2 - c)

### Associations between Pathway Polygenic Score and Mortality

*Analysis 3:* We investigated whether any of the three new “pathway-PGS” associate with time to mortality (7- Cluster - PGS, ChoMet - PGS, 6 - Clusters - PGS). We conducted the analyses first in the F2 nonagenarian siblings and then replicated the analysis in the L85+ sample.

*F2 nonagenarian siblings.* The analyses were conducted using random effect (frailty) Cox regression analysis:

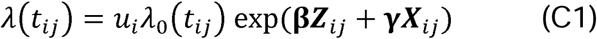

where t_ij_ is the age at death for person *j* in family *i*. λ_O_(t_ij_) refers to the baseline hazard, which is left unspecified. β is a vector of regression coefficients for the main effects of interest, representing the fixed effect of the variable z which correspond to a) the All - Clusters - PGS (C1 – a), b) the ChoMet_52_ – PGS (C1 – b), c) the 6 - Clusters - PGS (C1 – c), and d) the robustness check removing the SNPs that act on the *APOE* gene, the ChoMet_49_ – PGS (C1 – d). The models are adjusted for left truncation given by the age at entry in the study and sex (confounder) as denoted by x in expression 4. u_i_ > 0 refers to an unobserved random effect (frailty) shared by the members of the same family *i* and was assumed to follow a gamma distribution.

*L85+ sample.* The analyses were conducted using Cox regression analysis:

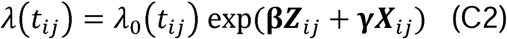

where t_ij_ is the age at death for person *j* in family *i*. λ_O_(t_ij_) refers to the baseline hazard, which is left unspecified. β is a vector of regression coefficients for the main effects of interest, representing the fixed effect of the variable z which corresponds to a) the All - Clusters - PGS (C2 – a), b) the ChoMet_52_ – PGS (C2 – b), and c) the 6 - Clusters - PGS (C1 – c), and d) the robustness check removing the SNPs that act on the *APOE* gene, the ChoMet_49_ – PGS (C1 – d). The models are adjusted for left truncation given by the age at entry in the study and confounders (sex and the four nearest genomic dimensions) as denoted by x in expression 5. The four nearest genomic dimensions were included to account for population stratification when performing analysis including PGS as depicted in Choi et al.^78^.

## Ethical declaration

Leiden Longevity Study and Leiden 85-plus Study: In accordance with the Declaration of Helsinki, we obtained informed consent from all participants prior entering the study. Good clinical practice guidelines were maintained. The study protocol was approved by the ethical committee of the Leiden University Medical Center before the start of the study (P01.113).

## Data availability

The individual-level data from the LLS and L85-plus are protected by Dutch personal integrity laws, and other (privacy) regulations. As such, restrictions apply to the availability of both cohort studies ‘data, which were used under license for the present work, thus are not publicly available. All summary statistics and data, underlying the main figures, are available in the source data file. For both datasets, additional summary statistics are available upon request to Pedro S.B. Ferreira (corresponding author; and Niels van den Berg (corresponding author). The LLS data is available following a data access procedure (https://leidenlangleven.nl/data-access/). Each request will be evaluated whether the research is compliant with the informed consent that has been signed by the LLS participants. The L85+ data is available for replication purposes upon request to Stella Trompet. Each request will be evaluated whether the research is compliant with the informed consent that has been signed by the L85+ participants and if replication is conducted within the secure Leiden University Medical Center network environment.

## Code availability

The scripts containing the code for data pre-processing, data analyses, and simulations performed in response to the reviewers can be freely downloaded at: https://git.lumc.nl/publications/familial_longevity_healthy_aging.

## Data Availability

The individual-level data from the LLS and L85-plus are protected by Dutch personal integrity laws, and other (privacy) regulations. As such, restrictions apply to the availability of both cohort studies data, which were used under license for the present work, thus are not publicly available. All summary statistics and data, underlying the main figures, are available in the source data file. For both datasets, additional summary statistics are available upon request to Pedro S.B. Ferreira (corresponding author; and Niels van den Berg (corresponding author). The LLS data is available following a data access procedure (https://leidenlangleven.nl/data-access/). Each request will be evaluated whether the research is compliant with the informed consent that has been signed by the LLS participants. The L85+ data is available for replication purposes upon request to Stella Trompet. Each request will be evaluated whether the research is compliant with the informed consent that has been signed by the L85+ participants and if replication is conducted within the secure Leiden University Medical Center network environment.

## Acknowledgments

The construction and maintenance of the LLS data has received funding from the European Union’s Seventh Framework Program (FP7/2007-2011) under grant agreement number 259679. This study was further supported by BBMRI-NL, a Research Infrastructure financed by the Dutch government (NWO 184.021.007 and 184.033.111). L85+ was funded in part by an unrestricted grant from the Dutch Ministry of Health, Welfare, and Sports. P.S.B.F. was funded by the Leiden University Fonds (Elise Mathilde foundation) and the Leiden University Medical Center’s Lifecourse Epidemiology and Geroscience theme grant. N.v.d.B. was funded by the Netherlands Organization for Scientific Research, domain Health Research and Medical Sciences (09120012010052). We thank Ryan Wan for dedicating his research masters’ internship to this topic.

## Author contribution

P. S. B. F. was responsible for data management, data analyses, writing the manuscript and finalizing it. N. v. d. B. conceived the project and provided supervisory input. M.B. provided overall project input and supervision. J.D. contributed to the analyses and methodology. P.S. provided overall project input. S.T. provided access to the Leiden 85-plus Study data and supported data organization. All authors contributed to the writing process leading to this paper.

## Supplementary Materials

See Supplementary Tables and Figures pdf file.

