## Supplementary Tables and Figures for "Low Genetic Risk for Coronary Artery Disease underlies Multigenerational Longevity and Healthy Aging"

**Table S1.** Robustness checks for association between LRC and CAD-PGS

| Method for Cardio-artery Disease PGS | Beta | P-value | N SNPs after QC |
|---|---|---|---|
| Genome-wide significant SNPs | -0.26 (-0.37, -0.15) | $3{,}68 \times 10^{-06}$ | 250 |
| Genome-wide significant SNPs minus APOE | -0.24 (-0.35, -0.13) | $2{,}23 \times 10^{-05}$ | 248 |
| Infinitesimal model (LDPred) | 0.01 (-0.09, -0.07) | $7{,}81 \times 10^{-01}$ | 2.324.683 |
| Genome-wide significant SNPs (FDR) | -0.22 (-0.33, -0.11) | $9{,}17 \times 10^{-05}$ | 897 |

Table shows the main analysis conducted with the Genome-wide significant SNPs for the Cardio-artery polygenic scores, the analysis removing APOE gene related SNPs (two), the main analysis using the infinitesimal model (significant and non-significant SNPs) based on LDPred, and the main analysis using FDR correction

**Table S2.** Association of LRC groups with Incidence of cardiovascular disease mediated by genetic predisposition to cardio artery disease

| | **Model 1** | | **Model 2** | | **Model 3** | |
|---|---|---|---|---|---|---|
| | **HR (95% CI)** | **P-value** | **HR (95% CI)** | **P-value** | **HR (95% CI)** | **P-value** |
| Coronary artery disease PGS | 1.41 (1.15-1.72) | $7.85 \times 10^{-04}$ | - | - | 1.33 (1.08-1.64) | $6.95 \times 10^{-03}$ |
| LRC30/LRC0 | - | - | 0.50 (0.33-0.78) | $1.99 \times 10^{-03}$ | 0.56 (0.36-0.86) | $8.43 \times 10^{-03}$ |
| Sex (Females) | 0.46 (0.32-0.67) | $4.42 \times 10^{-05}$ | 0.45 (0.30-0.67) | $7.35 \times 10^{-05}$ | 0.44 (0.30-0.66) | $5.50 \times 10^{-05}$ |
| Number of diseases pre-inclusion | 1.28 (0.96-1.70) | $8.96 \times 10^{-02}$ | 1.21 (0.90-1.64) | $2.13 \times 10^{-01}$ | 1.21 (0.89-1.64) | $2.17 \times 10^{-01}$ |
| Medication-use at inclusion | 1.19 (0.81-1.74) | $3.92 \times 10^{-01}$ | 1.25 (0.83-1.87) | $2.83 \times 10^{-01}$ | 1.23 (0.82-1.85) | $3.13 \times 10^{-01}$ |

*Note.* PGS = Polygenic Risk Score, LRC30/LRC0 = Comparison between Longevity Relatives' Count 30% group and Longevity Relatives Count 0% group

**Table S3**. Accelerated Failure Time Model

| | AFT1 | | AFT2 | | AFT3 | |
|---|---|---|---|---|---|---|
| | **TR (95% CI)** | **P-value** | **TR (95% CI)** | **P-value** | **TR (95% CI)** | **P-value** |
| Coronary artery disease PGS | 0.81 (0.72-0.92) | $1.35 \times 10^{-03}$ | - | - | 0.85 (0.75-0.96) | $7.95\times10^{-03}$ |
| LRC30/LRC0 | - | - | 1.54 (1.15-2.07) | $4.15 \times 10^{-03}$ | 1.41 (1.07-1.86) | $1.53\times10^{-02}$ |
| Sex (Females) | 1.67 (1.27-2.20) | $2.30 \times 10^{-04}$ | 1.70 (1.28-2.27) | $2.40 \times 10^{-04}$ | 1.68 (1.26-2.22) | $3.30\times10^{-04}$ |
| Number of diseases pre-inclusion | 0.85 (0.71-1.04) | $1.26 \times 10^{-01}$ | 0.89 (0.72-1.09) | $2.52 \times 10^{-01}$ | 0.90 (0.74-1.09) | $2.80\times10^{-01}$ |
| Medication-use at inclusion | 0.86 (0.66-1.12) | $2.65 \times 10^{-01}$ | 0.84 (0.63-1.11) | $2.29 \times 10^{-01}$ | 0.85 (0.65-1.12) | $2.59\times10^{-01}$ |
| Inclusion age | 0.96 (0.94-0.98) | $1.20 \times 10^{-05}$ | 0.96 (0.94-0.98) | $1.10 \times 10^{-04}$ | 0.96 (0.94-0.98) | $9.50\times10^{-05}$ |

Table shows the Accelerated Failure Time model for the coronary artery disease (CAD) Polygenic Score (PGS) and the incidence of cardiovascular disease (AFT1), the Accelerated Failure time Model for the LRC groups and incidence of cardiovascular disease (AFT2), and the mediation Accelerated Failure time Model (AFT3) based on the LRC groups associated to cardiovascular disease incidence mediated by the CAD-PGS. LRC refers to the longevity counts score and LRC30/LRC0 refers to the ratio between Longevity Relatives' Count 30% group and Longevity Relatives Count 0% group

**Table S4.** List of unique genes and respective SNPs of the CAD-PGS mapped from Aragam et al.

| rsID | Chromosome | Gene | ENSGID |
|---|---|---|---|
| rs10131894 | 14 | 75446879 | PGF |
| rs1034246 | 6 | 43068370 | VEGFA |
| rs10410487 | 19 | 17829608 | MYO9B |
| rs10488763 | 11 | 110244360 | RDX |
| rs10493891 | 16 | 81510742 | PLCG2 |
| rs1051338 | 10 | 91007360 | ACTA2 |
| rs10774625 | 12 | 111910219 | MYL2 |
| rs10951983 | 7 | 6446027 | RAC1 |
| rs11079536 | 17 | 62392403 | MAP3K3 |
| rs112949822 | 5 | 108085190 | FER |
| rs11466359 | 19 | 41837615 | TGFB1 |
| rs11523031 | 9 | 21843842 | CDKN2A |
| rs11601507 | 11 | 5701074 | ILK |
| rs11617955 | 13 | 110818102 | COL4A1 |
| rs11721038 | 3 | 168849576 | MECOM |
| rs11752218 | 6 | 57145562 | DST |
| rs1177562 | 11 | 118949331 | MCAM |
| rs12112877 | 7 | 106941324 | LAMB1 |
| rs1230666 | 1 | 114173410 | MAGI3 |
| rs1250247 | 2 | 216299629 | FN1 |
| rs12691049 | 16 | 15909513 | MYH11 |
| rs12740374 | 1 | 109817590 | CSF1 |
| rs13124853 | 4 | 146784774 | SMAD1 |
| rs13222797 | 7 | 117100046 | CAV1 |
| rs1510758 | 8 | 25061807 | DOCK5 |
| rs16952537 | 16 | 80185366 | WWOX |
| rs16986953 | 2 | 19942473 | RHOB |
| rs17080093 | 6 | 150997440 | AKAP12 |
| rs17083333 | 4 | 54572066 | PDGFRA |
| rs17086617 | 13 | 28962686 | FLT1 |
| rs17608766 | 17 | 45013271 | ITGB3 |
| rs17843797 | 3 | 124453022 | MYLK |
| rs185244 | 3 | 138092889 | MRAS |
| rs1870634 | 10 | 44480811 | CXCL12 |
| rs1892971 | 11 | 102795606 | PDGFD |
| rs2001846 | 8 | 126478450 | TRIB1 |
| rs2067831 | 10 | 105643223 | SH3PXD2A |
| rs2207132 | 20 | 39142516 | PLCG1 |
| rs2306363 | 11 | 65405600 | RELA |
| rs249760 | 5 | 141915692 | ARAP3 |
| rs2652682 | 5 | 17113657 | BASP1 |
| rs34759087 | 3 | 49162284 | LAMB2 |
| rs34991912 | 3 | 14926351 | FGD5 |
| rs357494 | 3 | 153937753 | ARHGEF26 |
| rs3918226 | 7 | 150690176 | NOS3 |
| rs4074793 | 5 | 52193125 | ITGA1 |
| rs429358 | 19 | 45411941 | APOE |
| rs4662330 | 2 | 144186475 | ZEB2 |
| rs476828 | 18 | 57852587 | LMAN1 |
| rs4790881 | 17 | 2068932 | PAFAH1B1 |
| rs4938809 | 11 | 120363937 | ARHGEF12 |
| rs55753709 | 10 | 96029170 | PLCE1 |
| rs56062135 | 15 | 67455630 | SMAD3 |
| rs582384 | 2 | 45896437 | EPAS1 |
| rs6026739 | 20 | 57739469 | GNAS |
| rs61797068 | 1 | 115902514 | NGF |

| | | | |
|---|---|---|---|
| rs633185 | 11 | 100593538 | ARHGAP42 |
| rs6686750 | 1 | 154419843 | IL6R |
| rs6909752 | 6 | 22612629 | PRL |
| rs6932293 | 6 | 160535878 | PLG |
| rs7077962 | 10 | 25054674 | ARHGAP21 |
| rs7133378 | 12 | 124409502 | UBC |
| rs71646019 | 1 | 59433354 | JUN |
| rs7183988 | 15 | 91428589 | FURIN |
| rs73025613 | 19 | 38334361 | ACTN4 |
| rs7500448 | 16 | 83045790 | CDH13 |
| rs7678555 | 4 | 120909501 | PDE5A |
| rs77347777 | 3 | 52848207 | NISCH |
| rs7903146 | 10 | 114758349 | TCF7L2 |
| rs79598313 | 1 | 27284913 | WASF2 |
| rs7991314 | 13 | 33126074 | STARD13 |
| rs8000794 | 13 | 99434810 | DOCK9 |
| rs8046696 | 16 | 75442143 | BCAR1 |
| rs8068844 | 17 | 40571284 | STAT3 |
| rs8176893 | 12 | 79999309 | PPP1R12A |
| rs9349379 | 6 | 12903957 | EDN1 |
| rs9400480 | 6 | 111850597 | LAMA4 |
| rs952227 | 2 | 227062080 | IRS1 |
| rs10176176 | 2 | 85762048 | GGCX |
| rs1019016 | 7 | 80570562 | CD36 |
| rs10477741 | 5 | 131795310 | AFF4 |
| rs10486389 | 7 | 20300416 | POLR1F |
| rs10790800 | 11 | 126262638 | ST3GAL4 |
| rs10811183 | 9 | 19436055 | DENND4C |
| rs10841443 | 12 | 20220033 | PDE3A |
| rs10852238 | 16 | 20253374 | DCUN1D3 |
| rs10857147 | 4 | 81181072 | ANTXR2 |
| rs10930115 | 2 | 164930382 | COBLL1 |
| rs10961206 | 9 | 13724051 | NFIB |
| rs11080107 | 17 | 27938424 | NEK8 |
| rs11107903 | 12 | 95507971 | FGD6 |
| rs111806192 | 16 | 28252382 | NUPR1 |
| rs11206803 | 1 | 56877509 | PLPP3 |
| rs112238647 | 15 | 79051705 | CTSH |
| rs112635299 | 14 | 94838142 | SERPINA1 |
| rs1132274 | 20 | 17596155 | RRBP1 |
| rs114192718 | 2 | 128785663 | LIMS2 |
| rs11556924 | 7 | 129663496 | UBE2H |
| rs11655024 | 17 | 59232365 | TBX2 |
| rs11663411 | 18 | 56960510 | ZNF532 |
| rs116843064 | 19 | 8429323 | ANGPTL4 |
| rs12445401 | 16 | 72148419 | ZFHX3 |
| rs12446515 | 16 | 56987015 | CETP |
| rs12468870 | 2 | 69679537 | AAK1 |
| rs12484557 | 22 | 24555861 | CABIN1 |
| rs12500824 | 4 | 77416627 | SEPTIN11 |
| rs12641981 | 4 | 45179883 | KCTD8 |
| rs12864131 | 13 | 27045939 | CDK8 |
| rs12916 | 5 | 74656539 | POLK |
| rs13120678 | 4 | 148273397 | EDNRA |
| rs13169691 | 5 | 118448279 | TNFAIP8 |
| rs139012 | 22 | 43623972 | PNPLA3 |
| rs1430158 | 2 | 183262128 | PDE1A |
| rs148812085 | 2 | 203877233 | SUMO1 |
| rs1536608 | 9 | 223613 | KANK1 |

| | | | |
|---|---|---|---|
| rs157333 | 5 | 156117200 | ADAM19 |
| rs16968377 | 17 | 36942396 | MED1 |
| rs17163363 | 1 | 222828704 | MIA3 |
| rs17263917 | 5 | 9552338 | SEMA5A |
| rs17566555 | 10 | 12275947 | CCDC3 |
| rs17581137 | 15 | 96146414 | NR2F2 |
| rs17680741 | 10 | 82251514 | ANXA11 |
| rs1807214 | 15 | 89565257 | MFGE8 |
| rs1967604 | 9 | 110530324 | ZNF462 |
| rs2008614 | 20 | 47433150 | B4GALT5 |
| rs2107595 | 7 | 19049388 | HDAC9 |
| rs2107732 | 7 | 45077978 | AEBP1 |
| rs215634 | 7 | 32369148 | PDE1C |
| rs2161967 | 2 | 218680529 | TNS1 |
| rs2215614 | 7 | 35277093 | TBX20 |
| rs2244608 | 12 | 121416988 | HNF1A |
| rs2277383 | 12 | 52314388 | ACVRL1 |
| rs2285219 | 1 | 175130983 | KIAA0040 |
| rs2410859 | 17 | 73841285 | SUMO2 |
| rs243071 | 2 | 60619028 | USP34 |
| rs2493298 | 1 | 3325912 | PRDM16 |
| rs2672592 | 10 | 124230750 | HTRA1 |
| rs268 | 8 | 19813529 | LPL |
| rs2681472 | 12 | 90008959 | ATP2B1 |
| rs2683260 | 15 | 81385552 | MESD |
| rs283485 | 2 | 233645691 | GIGYF2 |
| rs2843152 | 1 | 2245570 | SKI |
| rs28451064 | 21 | 35593827 | RUNX1 |
| rs2909217 | 17 | 66463985 | WIPI1 |
| rs2983896 | 6 | 97029871 | FHL5 |
| rs34232196 | 1 | 55489542 | PCSK9 |
| rs34330586 | 3 | 135800409 | STAG1 |
| rs34606058 | 12 | 115353368 | TBX3 |
| rs34917849 | 8 | 95278307 | GEM |
| rs35611688 | 2 | 148377860 | ACVR2A |
| rs360153 | 11 | 9762274 | IRAG1 |
| rs36033161 | 14 | 100123487 | CYP46A1 |
| rs3776307 | 5 | 142494165 | NR3C1 |
| rs3935875 | 9 | 139238824 | NOTCH1 |
| rs4140748 | 2 | 230005505 | PID1 |
| rs4245791 | 2 | 44074431 | ABCG5 |
| rs4266144 | 3 | 156852592 | TIPARP |
| rs4345341 | 5 | 121278751 | LOX |
| rs4452 | 22 | 33283257 | TIMP3 |
| rs4537761 | 11 | 9323353 | DENND2B |
| rs4643373 | 17 | 47123423 | PHB1 |
| rs4646249 | 8 | 18260431 | MTUS1 |
| rs4907571 | 13 | 113618496 | F10 |
| rs4954580 | 2 | 136986303 | R3HDM1 |
| rs515135 | 2 | 21286057 | APOB |
| rs55997232 | 19 | 11188117 | LDLR |
| rs56408342 | 8 | 22048490 | BMP1 |
| rs584961 | 11 | 75277628 | SERPINH1 |
| rs588136 | 15 | 58730498 | LIPC |
| rs6006426 | 22 | 30669883 | SMTN |
| rs60154123 | 1 | 210468999 | RCOR3 |
| rs6088595 | 20 | 33358499 | NCOA6 |
| rs61776719 | 1 | 38461319 | INPP5B |
| rs61806987 | 1 | 169314833 | ATP1B1 |

| | | | |
|---|---|---|---|
| rs62362364 | 5 | 55441571 | IL6ST |
| rs62405422 | 6 | 50796905 | TFAP2B |
| rs646668 | 10 | 116138034 | AFAP1L2 |
| rs651007 | 9 | 136153875 | CEL |
| rs6656344 | 1 | 42948585 | TIE1 |
| rs67807996 | 1 | 149995265 | ARNT |
| rs6883598 | 5 | 127926190 | FBN2 |
| rs6919211 | 6 | 133999868 | TCF21 |
| rs6953441 | 7 | 99617067 | GIGYF1 |
| rs7118294 | 11 | 32380521 | WT1 |
| rs71313931 | 22 | 19960184 | CLDN5 |
| rs7246865 | 19 | 17219105 | KLF2 |
| rs72836800 | 6 | 1617327 | FOXC1 |
| rs733701 | 6 | 39171862 | DAAM2 |
| rs7440763 | 4 | 156433520 | GUCY1A1 |
| rs75160195 | 12 | 54521594 | CBX5 |
| rs75655731 | 8 | 9721394 | TNKS |
| rs7622417 | 3 | 141625999 | ZBTB38 |
| rs77787671 | 10 | 104776205 | CYP17A1 |
| rs78030362 | 19 | 18575193 | GATAD2A |
| rs781663 | 4 | 57781754 | IGFBP7 |
| rs8075861 | 17 | 76915710 | C1QTNF1 |
| rs8124182 | 20 | 44608901 | PLTP |
| rs869396 | 4 | 169688000 | PALLD |
| rs884811 | 10 | 99923763 | LOXL4 |
| rs885150 | 9 | 124420173 | DAB2IP |
| rs9337951 | 10 | 30317073 | JCAD |
| rs9361867 | 6 | 82595959 | TENT5A |
| rs9399136 | 6 | 135402339 | AHI1 |
| rs9469899 | 6 | 34793124 | PPARD |
| rs964184 | 11 | 116648917 | APOA1 |
| rs9945890 | 18 | 46515916 | SMAD7 |
| rs9951447 | 18 | 20009691 | GATA6 |

**Table S5.** List of SNPs and Chromosomes included in the Pathway All-Clusters-PGS

| SNP | Chromosome | Gene |
|---|---|---|
| rs71646019 | 1 | JUN |
| rs61797068 | 1 | NGF |
| rs79598313 | 1 | WASF2 |
| rs6686750 | 1 | IL6R |
| rs1230666 | 1 | MAGI3 |
| rs11206803 | 1 | PLPP3 |
| rs11591147 | 1 | PCSK9 |
| rs12740374 | 1 | CSF1 |
| rs34232196 | 1 | PCSK9 |
| rs472495 | 1 | PCSK9 |
| rs56170783 | 1 | PLPP3 |
| rs61806987 | 1 | ATP1B1 |
| rs1250247 | 2 | FN1 |
| rs952227 | 2 | IRS1 |
| rs582384 | 2 | EPAS1 |
| rs6740731 | 2 | ZEB2 |
| rs10928241 | 2 | ZEB2 |
| rs4662330 | 2 | ZEB2 |
| rs16986953 | 2 | RHOB |
| rs4245791 | 2 | ABCG5 |
| rs515135 | 2 | APOB |
| rs76866386 | 2 | ABCG5 |
| rs185244 | 3 | MRAS |
| rs17843797 | 3 | MYLK |
| rs34759087 | 3 | LAMB2 |
| rs11721038 | 3 | MECOM |
| rs77347777 | 3 | NISCH |
| rs34991912 | 3 | FGD5 |
| rs17083333 | 4 | PDGFRA |
| rs13124853 | 4 | SMAD1 |
| rs7678555 | 4 | PDE5A |
| rs4074793 | 5 | ITGA1 |
| rs112949822 | 5 | FER |
| rs249760 | 5 | ARAP3 |
| rs2652682 | 5 | BASP1 |
| rs1034246 | 6 | VEGFA |
| rs6905288 | 6 | VEGFA |
| rs9400480 | 6 | LAMA4 |
| rs6909752 | 6 | PRL |
| rs9349379 | 6 | EDN1 |
| rs357494 | 6 | ARHGEF26 |
| rs11752218 | 6 | DST |
| rs17080093 | 6 | AKAP12 |
| rs10455872 | 6 | PLG |

| | | |
|---|---|---|
| rs186696265 | 6 | PLG |
| rs1998043 | 6 | PLG |
| rs6932293 | 6 | PLG |
| rs73596816 | 6 | PLG |
| rs9469899 | 6 | PPARD |
| rs10951983 | 7 | RAC1 |
| rs13222797 | 7 | CAV1 |
| rs12112877 | 7 | LAMB1 |
| rs1019016 | 7 | CD36 |
| rs3918226 | 7 | NOS3 |
| rs2001846 | 8 | TRIB1 |
| rs1510758 | 8 | DOCK5 |
| rs268 | 8 | LPL |
| rs56408342 | 8 | BMP1 |
| rs66778572 | 8 | LPL |
| rs894211 | 8 | LPL |
| rs2891168 | 9 | CDKN2A |
| rs11523031 | 9 | CDKN2A |
| rs76959412 | 9 | CDKN2A |
| rs6475608 | 9 | CDKN2A |
| rs651007 | 9 | CEL |
| rs1051338 | 10 | ACTA2 |
| rs7903146 | 10 | TCF7L2 |
| rs1870634 | 10 | CXCL12 |
| rs494207 | 10 | CXCL12 |
| rs55753709 | 10 | PLCE1 |
| rs7077962 | 10 | ARHGAP21 |
| rs2067831 | 10 | SH3PXD2A |
| rs2839812 | 11 | PDGFD |
| rs1892971 | 11 | PDGFD |
| rs4938809 | 11 | ARHGEF12 |
| rs633185 | 11 | ARHGAP42 |
| rs1177562 | 11 | MCAM |
| rs10488763 | 11 | RDX |
| rs11601507 | 11 | ILK |
| rs2306363 | 11 | RELA |
| rs964184 | 11 | APOA1 |
| rs8176893 | 12 | PPP1R12A |
| rs10774625 | 12 | MYL2 |
| rs2681472 | 12 | ATP2B1 |
| rs7133378 | 12 | UBC |
| rs7296737 | 12 | UBC |
| rs7485656 | 12 | UBC |
| rs17086617 | 13 | FLT1 |
| rs7991314 | 13 | STARD13 |
| rs8000794 | 13 | DOCK9 |
| rs11617955 | 13 | COL4A1 |

| | | |
|---|---|---|
| rs11619113 | 13 | COL4A1 |
| rs3783113 | 13 | COL4A1 |
| rs4773141 | 13 | COL4A1 |
| rs7333991 | 13 | COL4A1 |
| rs9515203 | 13 | COL4A1 |
| rs10131894 | 14 | PGF |
| rs112635299 | 14 | SERPINA1 |
| rs56062135 | 15 | SMAD3 |
| rs588136 | 15 | LIPC |
| rs7183988 | 15 | FURIN |
| rs8046696 | 16 | BCAR1 |
| rs12691049 | 16 | MYH11 |
| rs16952537 | 16 | WWOX |
| rs7500448 | 16 | CDH13 |
| rs7189462 | 16 | PLCG2 |
| rs10493891 | 16 | PLCG2 |
| rs12446515 | 16 | CETP |
| rs8068844 | 17 | STAT3 |
| rs17608766 | 17 | ITGB3 |
| rs11079536 | 17 | MAP3K3 |
| rs4790881 | 17 | PAFAH1B1 |
| rs4643373 | 17 | PHB1 |
| rs476828 | 18 | LMAN1 |
| rs11466359 | 19 | TGFB1 |
| rs1800469 | 19 | TGFB1 |
| rs73025613 | 19 | ACTN4 |
| rs10410487 | 19 | MYO9B |
| rs10422256 | 19 | LDLR |
| rs116843064 | 19 | ANGPTL4 |
| rs167479 | 19 | LDLR |
| rs429358 | 19 | APOE |
| rs55997232 | 19 | LDLR |
| rs7412 | 19 | APOE |
| rs8108474 | 19 | APOE |
| rs6102343 | 20 | PLCG1 |
| rs2207132 | 20 | PLCG1 |
| rs6026739 | 20 | GNAS |
| rs8124182 | 20 | PLTP |
| rs149487184 | 21 | RUNX1 |
| rs28451064 | 21 | RUNX1 |
| rs4452 | 22 | TIMP3 |

**Table S6.** List of SNPs and Chromosomes included in the Pathway $ChoMet_{52}$-PGS

| SNP | Chromosome | Gene |
|---|---|---|
| rs61806987 | 1 | ATP1B1 |
| rs12740374 | 1 | CSF1 |
| rs11591147 | 1 | PCSK9 |
| rs34232196 | 1 | PCSK9 |
| rs472495 | 1 | PCSK9 |
| rs11206803 | 1 | PLPP3 |
| rs56170783 | 1 | PLPP3 |
| rs4245791 | 2 | ABCG5 |
| rs76866386 | 2 | ABCG5 |
| rs515135 | 2 | APOB |
| rs10455872 | 6 | PLG |
| rs186696265 | 6 | PLG |
| rs1998043 | 6 | PLG |
| rs6932293 | 6 | PLG |
| rs73596816 | 6 | PLG |
| rs9469899 | 6 | PPARD |
| rs1019016 | 7 | CD36 |
| rs3918226 | 7 | NOS3 |
| rs56408342 | 8 | BMP1 |
| rs268 | 8 | LPL |
| rs66778572 | 8 | LPL |
| rs894211 | 8 | LPL |
| rs651007 | 9 | CEL |
| rs964184 | 11 | APOA1 |
| rs11601507 | 11 | ILK |
| rs10488763 | 11 | RDX |
| rs2306363 | 11 | RELA |
| rs2681472 | 12 | ATP2B1 |
| rs7133378 | 12 | UBC |
| rs7296737 | 12 | UBC |
| rs7485656 | 12 | UBC |
| rs11617955 | 13 | COL4A1 |
| rs11619113 | 13 | COL4A1 |
| rs3783113 | 13 | COL4A1 |
| rs4773141 | 13 | COL4A1 |
| rs7333991 | 13 | COL4A1 |
| rs9515203 | 13 | COL4A1 |
| rs7183988 | 15 | FURIN |
| rs588136 | 15 | LIPC |
| rs12446515 | 16 | CETP |
| rs4643373 | 17 | PHB1 |
| rs116843064 | 19 | ANGPTL4 |
| rs429358 | 19 | APOE |

| | | |
|---|---|---|
| rs7412 | 19 | APOE |
| rs8108474 | 19 | APOE |
| rs10422256 | 19 | LDLR |
| rs167479 | 19 | LDLR |
| rs55997232 | 19 | LDLR |
| rs6026739 | 20 | GNAS |
| rs8124182 | 20 | PLTP |
| rs149487184 | 21 | RUNX1 |
| rs28451064 | 21 | RUNX1 |

**Table S7.** List of SNPs and Chromosomes included in the Pathway 6-Clusters-PGS

| SNP | Chromosome | Gene |
|---|---|---|
| rs71646019 | 1 | JUN |
| rs61797068 | 1 | NGF |
| rs79598313 | 1 | WASF2 |
| rs6686750 | 1 | IL6R |
| rs1230666 | 1 | MAGI3 |
| rs1250247 | 2 | FN1 |
| rs952227 | 2 | IRS1 |
| rs582384 | 2 | EPAS1 |
| rs6740731 | 2 | ZEB2 |
| rs10928241 | 2 | ZEB2 |
| rs4662330 | 2 | ZEB2 |
| rs16986953 | 2 | RHOB |
| rs185244 | 3 | MRAS |
| rs17843797 | 3 | MYLK |
| rs34759087 | 3 | LAMB2 |
| rs11721038 | 3 | MECOM |
| rs77347777 | 3 | NISCH |
| rs34991912 | 3 | FGD5 |
| rs17083333 | 4 | PDGFRA |
| rs13124853 | 4 | SMAD1 |
| rs7678555 | 4 | PDE5A |
| rs4074793 | 5 | ITGA1 |
| rs112949822 | 5 | FER |
| rs249760 | 5 | ARAP3 |
| rs2652682 | 5 | BASP1 |
| rs1034246 | 6 | VEGFA |
| rs6905288 | 6 | VEGFA |
| rs9400480 | 6 | LAMA4 |
| rs6909752 | 6 | PRL |
| rs9349379 | 6 | EDN1 |
| rs357494 | 6 | ARHGEF26 |
| rs11752218 | 6 | DST |
| rs17080093 | 6 | AKAP12 |
| rs10951983 | 7 | RAC1 |
| rs13222797 | 7 | CAV1 |
| rs12112877 | 7 | LAMB1 |
| rs2001846 | 8 | TRIB1 |
| rs1510758 | 8 | DOCK5 |
| rs2891168 | 9 | CDKN2A |
| rs11523031 | 9 | CDKN2A |
| rs76959412 | 9 | CDKN2A |
| rs6475608 | 9 | CDKN2A |
| rs1051338 | 10 | ACTA2 |
| rs7903146 | 10 | TCF7L2 |

| | | |
|---|---|---|
| rs1870634 | 10 | CXCL12 |
| rs494207 | 10 | CXCL12 |
| rs55753709 | 10 | PLCE1 |
| rs7077962 | 10 | ARHGAP21 |
| rs2067831 | 10 | SH3PXD2A |
| rs2839812 | 11 | PDGFD |
| rs1892971 | 11 | PDGFD |
| rs4938809 | 11 | ARHGEF12 |
| rs633185 | 11 | ARHGAP42 |
| rs1177562 | 11 | MCAM |
| rs8176893 | 12 | PPP1R12A |
| rs10774625 | 12 | MYL2 |
| rs17086617 | 13 | FLT1 |
| rs7991314 | 13 | STARD13 |
| rs8000794 | 13 | DOCK9 |
| rs10131894 | 14 | PGF |
| rs112635299 | 14 | SERPINA1 |
| rs56062135 | 15 | SMAD3 |
| rs8046696 | 16 | BCAR1 |
| rs12691049 | 16 | MYH11 |
| rs16952537 | 16 | WWOX |
| rs7500448 | 16 | CDH13 |
| rs7189462 | 16 | PLCG2 |
| rs10493891 | 16 | PLCG2 |
| rs8068844 | 17 | STAT3 |
| rs17608766 | 17 | ITGB3 |
| rs11079536 | 17 | MAP3K3 |
| rs4790881 | 17 | PAFAH1B1 |
| rs476828 | 18 | LMAN1 |
| rs11466359 | 19 | TGFB1 |
| rs1800469 | 19 | TGFB1 |
| rs73025613 | 19 | ACTN4 |
| rs10410487 | 19 | MYO9B |
| rs6102343 | 20 | PLCG1 |
| rs2207132 | 20 | PLCG1 |
| rs4452 | 22 | TIMP3 |

**Table S8.** Association between Pathway-PGSs and All-Cause-Mortality

| Cohort / Regression Method | All-Clusters-PGS | | $ChoMet_{52}$-PGS | | 6-Clusters-PGS | |
|---|---|---|---|---|---|---|
| | **HR (95% CI)** | **P-value** | **HR (95% CI)** | **P-value** | **HR (95% CI)** | **P-value** |
| **Panel A: Leiden Longevity Study F2 generation (Frailty Cox)** | | | | | | |
| PGS | 1.08 (1.00-1.16) | $5.00 \times 10^{-02}$ | 1.12 (1.04-1.20) | $2.62 \times 10^{-03}$ | 0.98 (0.91-1.05) | $4.78 \times 10^{-01}$ |
| Sex (Females) | 0.73 (0.63-0.85) | $7.05 \times 10^{-05}$ | 0.74 (0.63-0.86) | $8.71 \times 10^{-05}$ | 0.73 (0.62-0.85) | $5.96 \times 10^{-05}$ |
| Random effect variance | 0.10 | | 0.09 | | 0.10 | |
| **Panel B: Leiden 85-plus Study (Cox)** | | | | | | |
| PGS | 1.08 (0.97 - 1.20) | $1.18 \times 10^{-01}$ | 1.11 (1.00 – 1.23) | $4.93 \times 10^{-02}$ | 1.01 (0.90 - 1.13) | $9.15 \times 10^{-01}$ |
| Sex (Females) | 0.71 (0.56 - 0.92) | $7.94 \times 10^{-03}$ | 0.72 (0.56 – 0.92) | $8.34 \times 10^{-03}$ | 0.71 (0.53 - 0.91) | $6.73 \times 10^{-03}$ |

**Table S9.** *Demographic characteristics of the Leiden Longevity Study (LLS) ancestral groups used to estimate the LRC Score*

| | **Ancestral family groups: F3 children** | | | **Ancestral family groups: F3 partners** |
|---|---|---|---|---|
| | **F1 grandparents of the children** | **F2 parents of the children** | **F2 aunts and uncles** | **F2 parents of the partners** |
| Number, N (number of families) | 760 | 1295 | 2364 | 1252 |
| Female, N (%) | 380 (50) | 646 (50) | 1167 (49) | 626 (50) |
| Range birth cohorts, years | 1850-1894 | 1882-1928 | 1875-1941 | 1864 - 1947 |
| Alive, N (%) | 0 (0) | 22 (2) | 361 (15) | 222 (18) |
| Deceased, N (%) | 760 (100) | 1179 (91) | 1974 (84) | 1010 (81) |
| Missing age, N (%) | 0 (0) | 94 (7) | 33 (1) | 37 (3) |

*Note.* For all groups, mortality information was updated in January 2021. All mortality information was obtained from the official and verified Netherlands Population Registers

**Table S10.** DNA isolation and Genotyping overview for the Leiden Longevity Study

| Study | Ethnicity | Country | Genotyping Array | Genotyping calling algorithm | Sample call rate | Other exclusions | MAF (required) | HWE (required) | CALL RATE (required) | Mendelian Error (exclude if) | Duplicate Errors (exclude if) | Other | SNPs for imputation | Reference Panel | Build | Software for imputation | Filters | SNPs for analysis (imputed and genotyped) | Reference study description (PMID) | Study link/ website |
|---|---|---|---|---|---|---|---|---|---|---|---|---|---|---|---|---|---|---|---|---|
| Leiden Longevity Study | European | Netherlands | Illumina Human660W and OmniExpress | Illumina GenomeStudio | >95% | NA | >= 1% | $>=10^{-4}$ | >= 95% | NA | NA | NA | 296,486 autosomal and 6,185 X-linked | HRC | 37 | HRC server | NA | 39,117,106 | 19682117 | www.leidenlanglev en.nl/en/home |

From all participants in the Leiden Longevity Study, blood was drawn in a 10-ml coagulation Vacutainer (BD Alphen a/d Rijn), 8-ml and 4.5-ml EDTA Vacutainers, an 8-ml citrate Vacutainer, and a 2.5-ml PAXgene tube (PreAnalytiX, Hombrechtikon). Serum, EDTA, and citrate plasma were stored at −80°C, and EDTA and citrate buffy coats at −20°C. DNA was isolated from the EDTA buffy coats. From generation 2, the whole buffy coat was isolated using a QIAamp DNA Blood Maxi Kit (Qiagen, Venlo) according to the manufacturer's protocol. DNA isolation from generation 3 was outsourced to BaseClear (Leiden, The Netherlands), where 200 µl of the buffy coat was isolated using the Chemagic DNA Blood 100 Kit (Chemagen, Baesweiler) and carried out according to the manufacturer's protocol on a Magnetic Separation Module I (Chemagen), equipped with a 96-rod head. All DNA concentrations were determined using OD260 measurement.

**Table S11.** DNA isolation and Genotyping overview of Leiden 85-plus Study

| Study | Ethnicity | Country | Genotyping Array | Genotyping calling algorithm | Sample call rate | Other exclusions | MAF (required) | HWE (required) | CALL RATE (required) | Mendelian Error (exclude if) | Duplicate Errors (exclude if) | Other | SNPs for imputation | Reference Panel | Build | Software for imputation | Filters | SNPs for analysis (imputed and genotyped) | Reference study description (PMID) | Study link/ website |
|---|---|---|---|---|---|---|---|---|---|---|---|---|---|---|---|---|---|---|---|---|
| Leiden 85-plus Study | European | Netherlands | Illumina Human660W and OmniExpress | Illumina Genome Studio | >95% | NA | >= 1% | P>=$10^{-4}$ | >= 95% | NA | NA | NA | 601,010 autosomal | HRC | 37 | Michigan Imputation Server 2 | $R^2$-filter = 0.3 | 13,188,172 | 25855712 | https://doi.org/10.1093/ije/dyv031 |

In the Leiden 85-plus, 335 individuals were included for imputation. Quality control after imputation removed SNPs with MAF ≥ 1%, HWE ≥ $10^{-6}$ , geno ≥ 0.05, missingness ≥ 0.05, prunning with a window size of 200 variants with step size of 50 variants at a time, and filtering out SNPs with LD r2 higher than 0.25. In addition, individuals with PIHAT ≥ 0.125 were excluded, leaving a final sample of 319 individuals and 236,609 SNPs for analyses.

**Table S12.** Disease Domains, Specific Diseases, and matched Genome Wide Association Studies (GWAS)

| Disease Domain | Disease Specific Cause of Deaht | Percentage of Deaths within Domain | Rank | GWAS | Paper DOI |
|---|---|---|---|---|---|
| Neoplasms | C34 Malignant neoplasm of bronchus and lung | 21% | 1 | Lung Cancer | https://doi.org/10.1038/s41588-022-01115-x |
| (C00-D48) | C18 Malignant neoplasm of colon | 7% | 2 | Colon Cancer | https://doi.org/10.1038/s41588-022-01222-9 |
| | C50 Malignant neoplasm of breast | 7% | 3 | Breast Cancer | https://doi.org/10.1038/s41467-020-15046-w |
| | C61 Malignant neoplasm of prostate | 7% | 4 | Prostate Cancer | https://doi.org/10.1038/s41588-020-00748-0 |
| | C25 Malignant neoplasm of pancreas | 6% | 5 | Pancreatic cancer | https://doi.org/10.1038/s41467-018-02942-5 |
| Circulatory System | I50 Heart failure | 20% | 1 | Coronary artery disease | https://doi.org/10.1038/s41588-022-01233-6 |
| (I00-I99) | I21 Acute myocardial infarction | 10% | 2 | Myocardial infarction | https://doi.org/10.1093/eurheartj/ehaa1040 |
| | I64 Stroke, not specified as haemorrhage or infarction | 4% | 3 | Stroke | https://doi.org/10.1038/s41586-022-05165-3 |
| | I63 Cerebral infarction | 4% | 4 | Stroke | https://doi.org/10.1038/s41586-022-05165-3 |
| | I61 Intracerebral haemorrhage | 4% | 5 | Stroke | https://doi.org/10.1038/s41586-022-05165-3 |
| Psychological Disorders | F03 Unspecified dementia | 79% | 1 | Dementia | https://doi.org/10.1016/j.archger.2022.104853 |
| (F00-F99) | F01 Vascular dementia | 10% | 2 | N/A | - |
| | F10 Mental disorders due to alcohol | 5% | 3 | N/A | - |
| | F32 Depressive episode | 1% | 4 | Major depressive disorder | https://doi.org/10.1038/s41593-018-0326-7 |
| | F05 Delirium, not induced by alcohol and other psychoactive substances | 1% | 5 | N/A | - |
| Respiratory System | J44 Other chronic obstructive pulmonary disease | 53% | 1 | COPD | https://doi.org/10.1038/s41588-018-0342-2 |
| (J00-J99) | J18 Pneumonia, organism unspecified | 25% | 2 | N/A | - |
| | J84 Other interstitial pulmonary disease | 7% | 3 | N/A | - |
| | J69 Pneumonitis due to solids and liquids | 3% | 4 | N/A | - |
| | J98 Other respiratory disorders | 2% | 5 | N/A | - |
| Nervous System | G30 Alzheimer's disease | 43% | 1 | Alzheimer's disease | https://doi.org/10.1038/s41588-022-01024-z |
| (G00-G99) | G20 Parkinson's disease | 22% | 2 | Parkingson's disease | https://doi.org/10.1016/s1474-4422(19)30320-5 |
| | G12 Spinal muscular atrophy and related syndromes | 7% | 3 | N/A | - |
| | G31 Other degenerative diseases of nervous system, not elsewhere classified | 7% | 4 | N/A | - |
| | G35 Multiple sclerosis | 3% | 5 | N/A | - |
| Digestive System | K92 Other diseases of digestive system | 17% | 1 | N/A | - |
| (K00-K93) | K56 Paralytic ileus and intestinal obstruction without hernia | 13% | 2 | N/A | - |
| | K74 Fibrosis and cirrhosis of liver | 10% | 3 | N/A | - |
| | K55 Vascular disorders of intestine | 9% | 4 | N/A | - |
| | K70 Alcoholic liver disease | 8% | 5 | N/A | - |
| Endocrine, Nutritional, and Metabolic Diseases | E14 Unspecified diabetes mellitus | 47% | 1 | Type 2 diabetes | https://doi.org/10.1038/s41588-022-01058-3 |
| (E00-E90) | E11 Non-insulin-dependent diabetes mellitus | 25% | 2 | Type 2 diabetes | https://doi.org/10.1038/s41588-022-01058-3 |
| | E66 Obesity | 6% | 3 | N/A | - |
| | E10 Insulin-dependent diabetes mellitus | 5% | 4 | N/A | - |
| | E85 Amyloidosis | 5% | 5 | N/A | - |
| Genitourinary System | N39 Other disorders of urinary system | 33% | 1 | N/A | - |
| (N00-N99) | N18 Chronic renal failure | 29% | 2 | Renal failure | https://doi.org/10.1016/j.kint.2022.05.021 |
| | N19 Unspecified renal failure | 18% | 3 | Renal failure | https://doi.org/10.1016/j.kint.2022.05.021 |
| | N17 Acute renal failure | 7% | 4 | Renal failure | https://doi.org/10.1016/j.kint.2022.05.021 |
| | N28 Other disorders of kidney and ureter, not elsewhere classified | 2% | 5 | N/A | - |
| Infectious Diseases | A41 Other septicaemia | 33% | 1 | Bacterial/viral susceptibility | https://doi.org/10.1038/s41598-022-05838-z |
| (A00-B99) | B99 Other and unspecified infectious diseases | 26% | 2 | Bacterial/viral susceptibility | https://doi.org/10.1038/s41598-022-05838-z |
| | A09 Diarrhoea and gastro-enteritis of presumed infectious origin | 12% | 3 | Bacterial/viral susceptibility | https://doi.org/10.1038/s41598-022-05838-z |
| | A49 Bacterial infection of unspecified site | 6% | 4 | Bacterial/viral susceptibility | https://doi.org/10.1038/s41598-022-05838-z |
| | A46 Erysipelas | 5% | 5 | N/A | - |
| Musculoskeletal System | M06 Other rheumatoid arthritis | 14% | 1 | Rheumatoid arthritis | https://doi.org/10.1038/s41588-022-01213-w |
| (M00-M99) | M16 Coxarthrosis [arthrosis of hip] | 7% | 2 | Osteoarthritis | https://doi.org/10.1016/j.cell.2021.07.038 |
| | M48 Other spondylopathies | 7% | 3 | Osteoarthritis | https://doi.org/10.1016/j.cell.2021.07.038 |
| | M35 Other systemic involvement of connective tissue | 6% | 4 | N/A | - |
| | M31 Other necrotising vasculopathies | 5% | 5 | N/A | - |

**Table S13**: Disease Domains, Disease-PGS, and QC information

| Disease Domain | Disease-PGS | N SNPs before QC | N SNPs after QC | Reference |
|---|---|---|---|---|
| Neoplasms | Breast Cancer | 31 | 23 | https://doi.org/10.1038/s41467-020-15046-w |
| Neoplasms | Colorectal Cancer | 205 | 193 | https://doi.org/10.1038/s41588-022-01222-9 |
| Neoplasms | Lung Cancer | 45 | 33 | https://doi.org/10.1038/s41588-022-01115-x |
| Neoplasms | Pancreatic Cancer | 32 | 27 | https://doi.org/10.1038/s41467-018-02942-5 |
| Neoplasms | Prostate Cancer | 268 | 209 | https://doi.org/10.1038/s41588-020-00748-0 |
| Circulatory System | Coronary Artery Disease | 279 | 250 | https://doi.org/10.1038/s41588-022-01233-6 |
| Circulatory System | Myocardial Infarction | 213 | 197 | https://doi.org/10.1093/eurheartj/ehaa1040 |
| Circulatory System | Stroke | 17 | 17 | https://doi.org/10.1038/s41586-022-05165-3 |
| Psychological Disorders | Dementia | 27 | 26 | https://doi.org/10.1016/j.archger.2022.104853 |
| Psychological Disorders | Depression | 102 | 95 | https://doi.org/10.1038/s41593-018-0326-7 |
| Respiratory System | COPD | 82 | 79 | https://doi.org/10.1038/s41588-018-0342-2 |
| Nervous System | Alzheimer’s Disease | 83 | 72 | https://doi.org/10.1038/s41588-022-01024-z |
| Nervous System | Parkinson’s Disease | 38 | 37 | https://doi.org/10.1016/s1474-4422(19)30320-5 |
| Endocrine, Nutritional, and Metabolic | Type 2 Diabetes | 338 | 318 | https://doi.org/10.1038/s41588-022-01058-3 |
| Genitourimary System | Renal Failure | 12 | 10 | https://doi.org/10.1016/j.kint.2022.05.021 |
| Infectious Susceptibility | Susceptibility to Bacterial and Viral Infections | 57 | 21 | https://doi.org/10.1038/s41598-022-05838-z |
| Musculoskeletal System | Osteoarthritis | 13 | 9 | https://doi.org/10.1016/j.cell.2021.07.038 |
| Musculoskeletal System | Rheumatoid Arthritis | 123 | 95 | https://doi.org/10.1038/s41588-022-01213-w |

**Table S14.** Comparison among different distributions to fit Accelerated Failure Time Model

| **Model** | **AIC** | **BIC** |
|---|---|---|
| Exponential | 1492,618 | 1523,82 |
| Weibull | 1480,137 | 1516,54 |
| Gaussian | 1494,445 | 1530,848 |
| Logistic | 1506,785 | 1543,188 |
| **Log-logistic** | **1479,563** | **1515,966** |
| Log-normal | 1489,461 | 1525,864 |

Table shows the best fitting distribution for using Accelerated Failure Time Model (parametric model). Best fitting model highlighted in bold

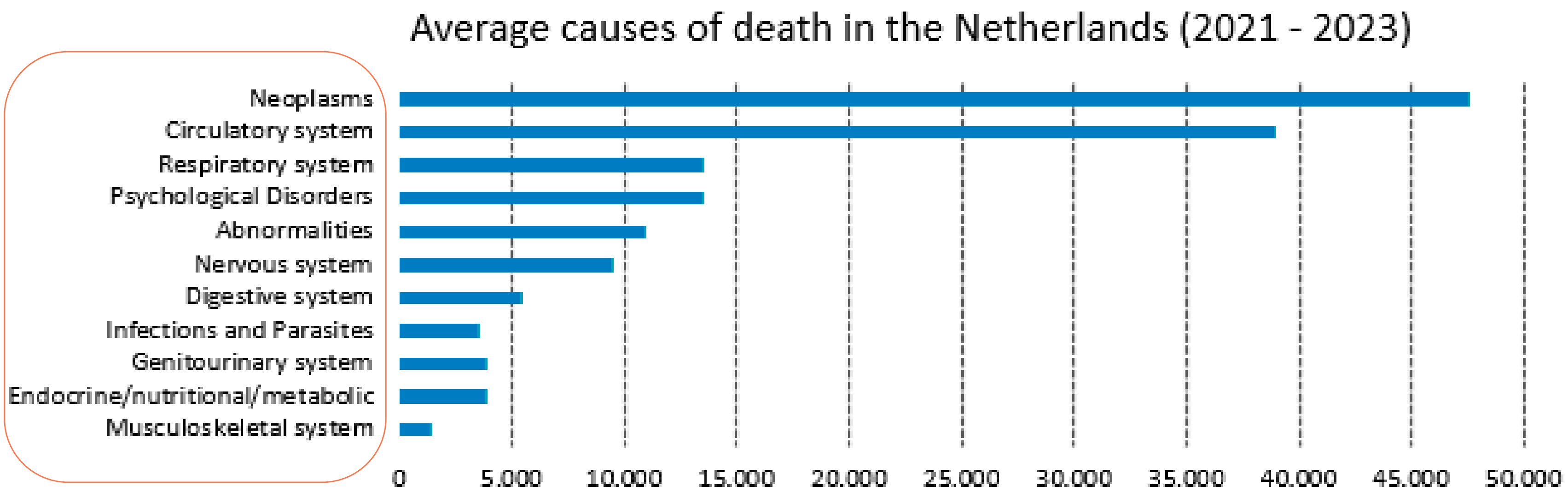


**Figure S1. | Average number of deaths in The Netherlands between 2021 and 2023 by disease domains**
Figure shows the average of deaths in The Netherlands per disease domain according to the Dutch Statistics Bureau (CBS). Deaths do to the COVID-19 were excluded.

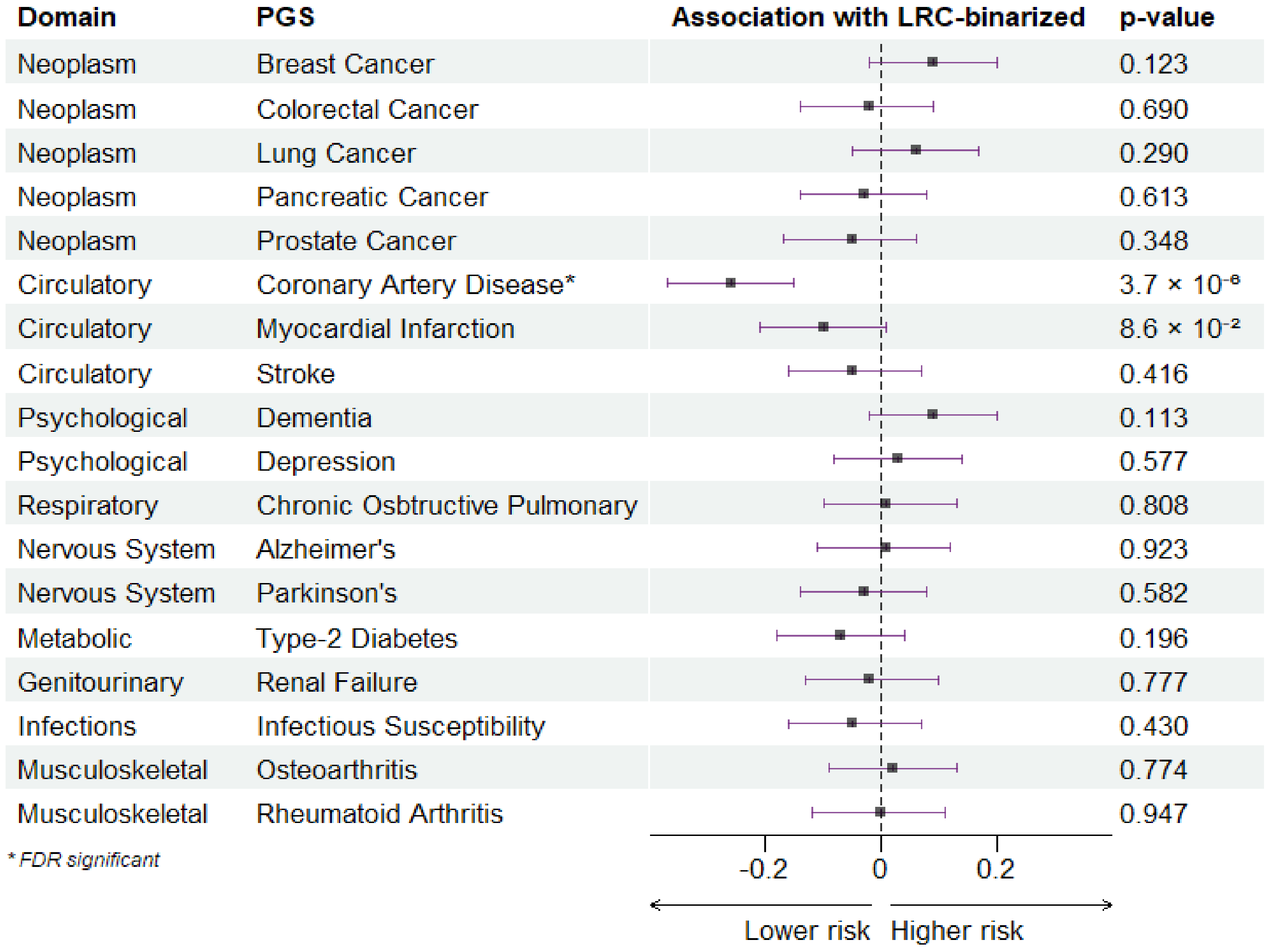


**Figure S2.** | **Association between LRC Groups (binarized) and Polygenic Scores for diseases.**

The figure shows the association between the Longevity Relatives Count score groups (LRC-groups) and the Polygenic Scores (PGS) for the top 18 diseases causes of death in the Netherlands according to the Dutch Central Statistics Bureau (CBS). Negative values indicate that LRC30% have a lower genetic risk to the disease in comparison to LRC0%. The asterisk indicates which associations were significant after FDR correction for multiple testing.

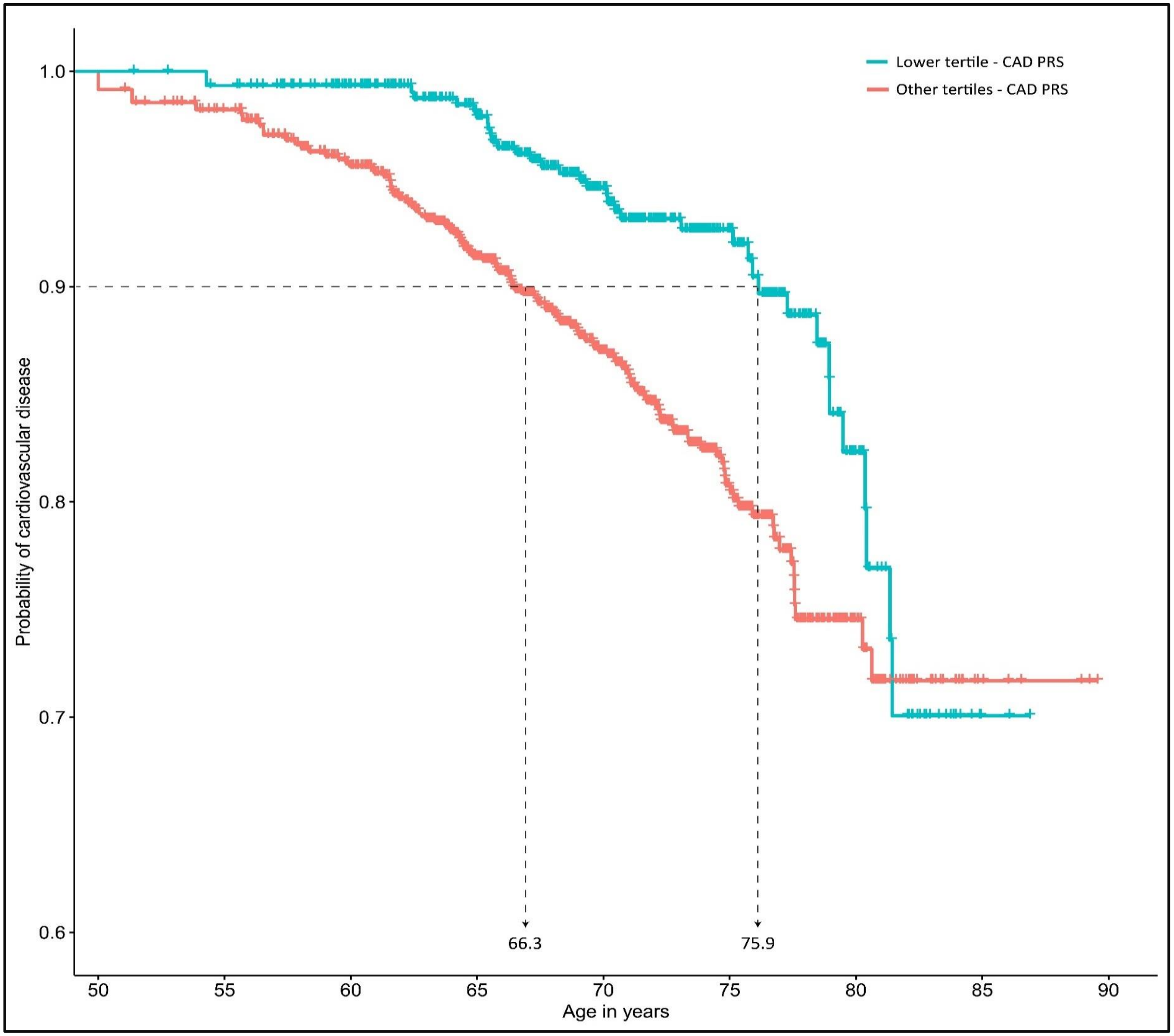


**Figure S3 | Individuals in the lower tertile for the CAD-PGS show delayed CVD onset**

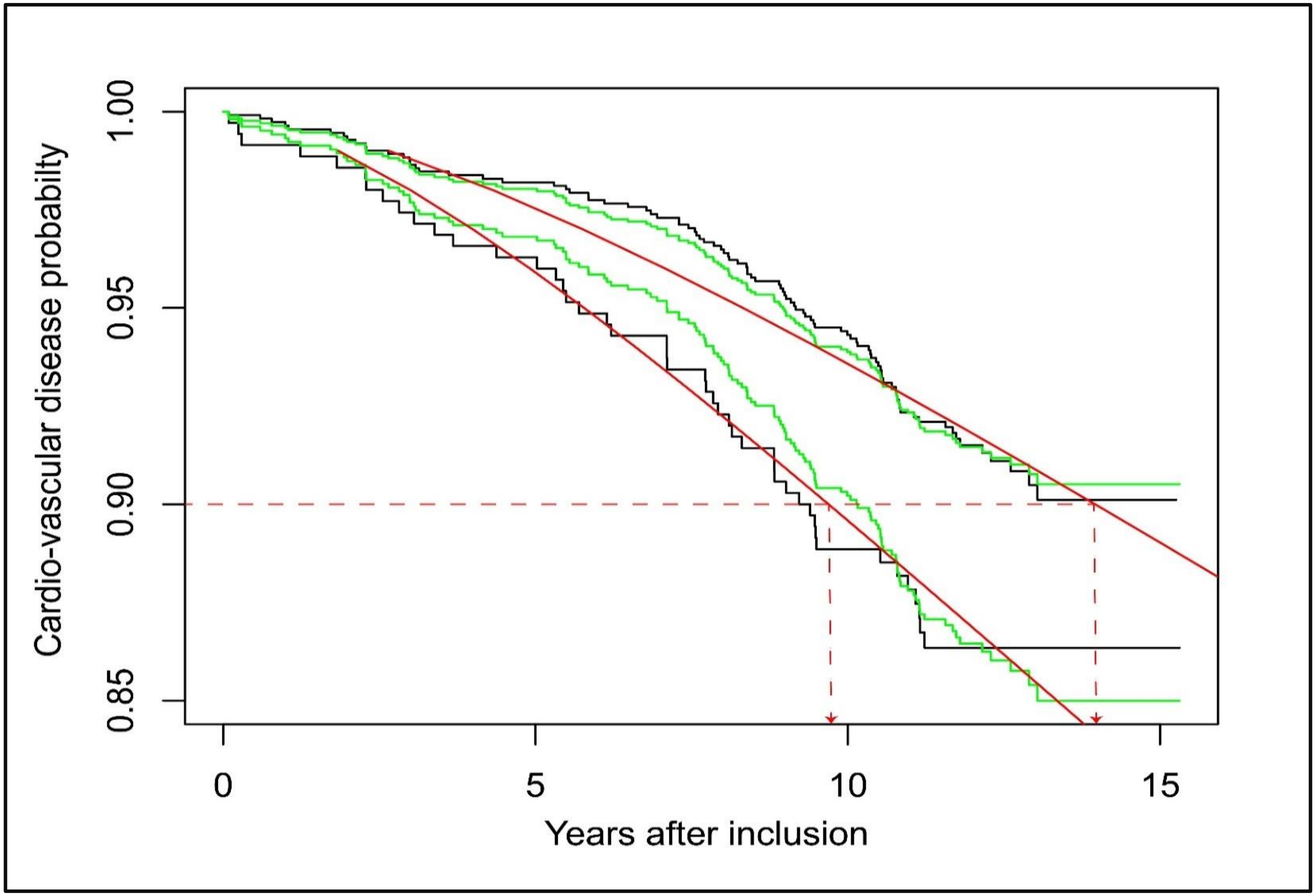


**Figure S4 | Overlap between Frailty Models and AFT Models shows delayed CVD onset for members of long-lived families**

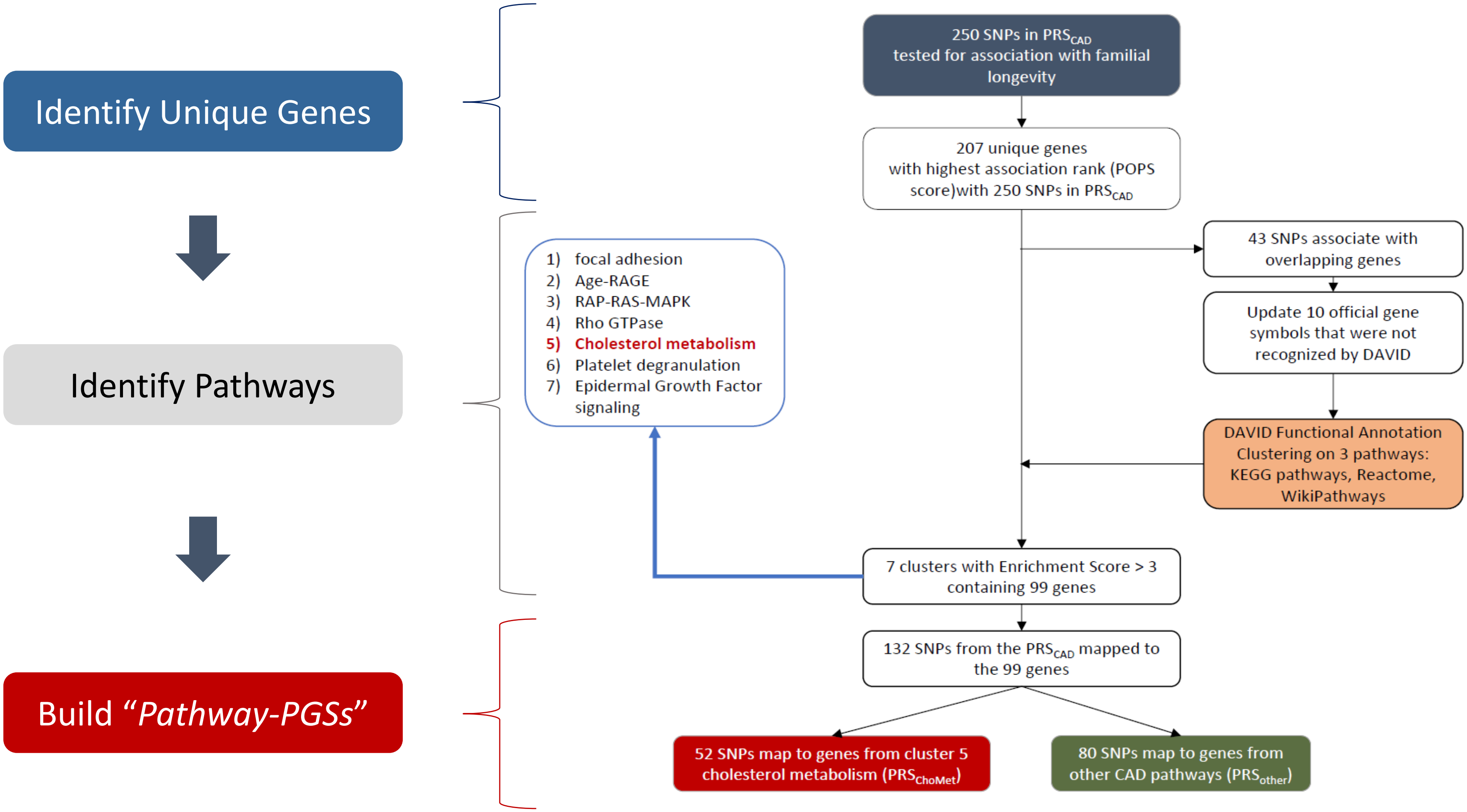


**Figure S5. | Overview of the procedure to construct PGSs based on pathways indentified with DAVID**

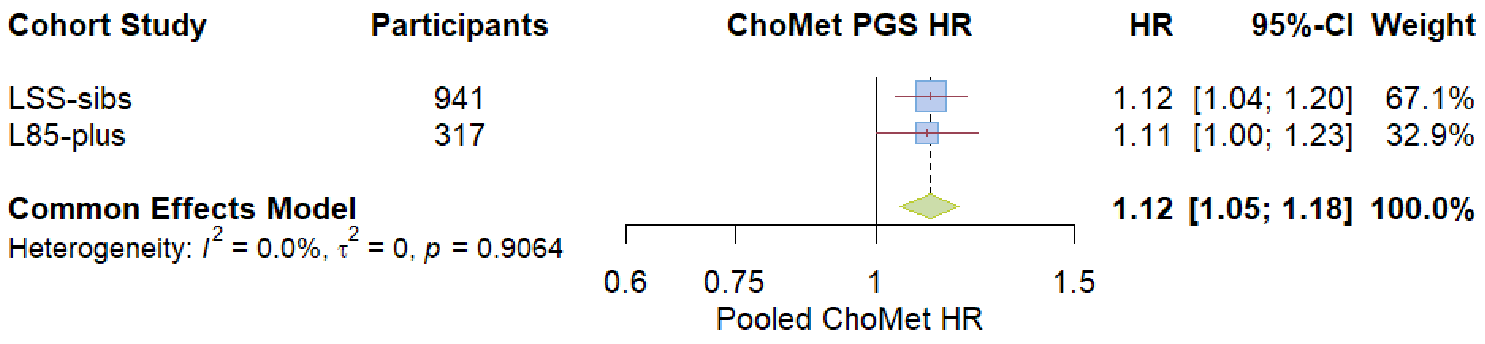


**Figure S6.** | **Meta-analysis points to increased risk of dying with an increase in the ChoMet$_{52}$-PGS**

The Figure shows a pooled effect for the ChoMet$_{52}$-PGS based on a common (fixed) effects meta-analysis with the effects observed for the ChoMet$_{52}$-PGS in the Leiden Longevity Study and the Leiden 85-plus study. Blue squares represent the weight of each cohort study. Red line represents the 95% confidence interval. Green Diamond represents the pooled effect with 95% confidence interval. With one standard deviation increase in the pooled effect for the ChoMet$_{52}$-PGS, the risk of dying increases 12%.

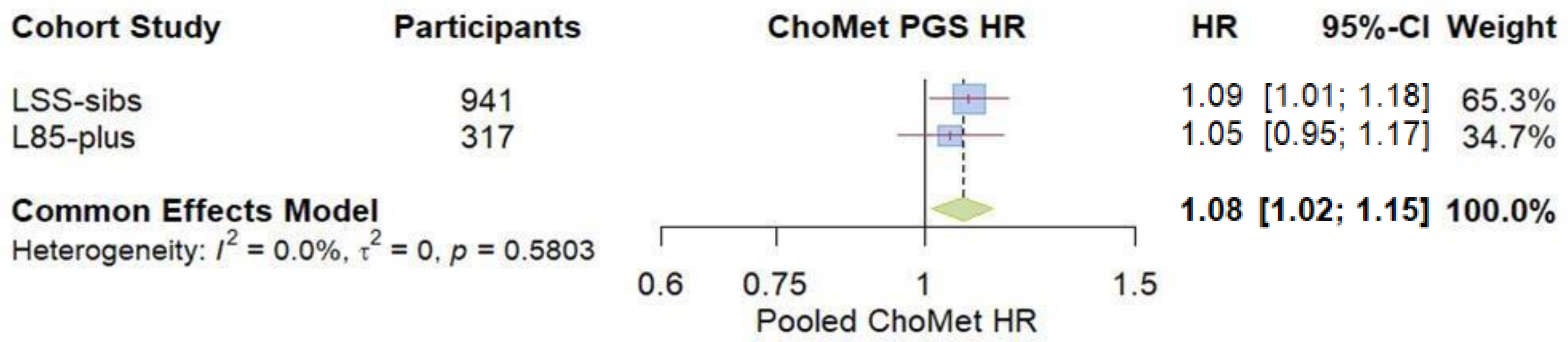


**Figure S7. | Meta-Analysis points to increased risk of dying with an increase in the ChoMet$_{49}$-PGS**

Figure shows that the pooled effect for the ChoMet$_{49}$-PGS, which does not include APOE SNPs, is significant

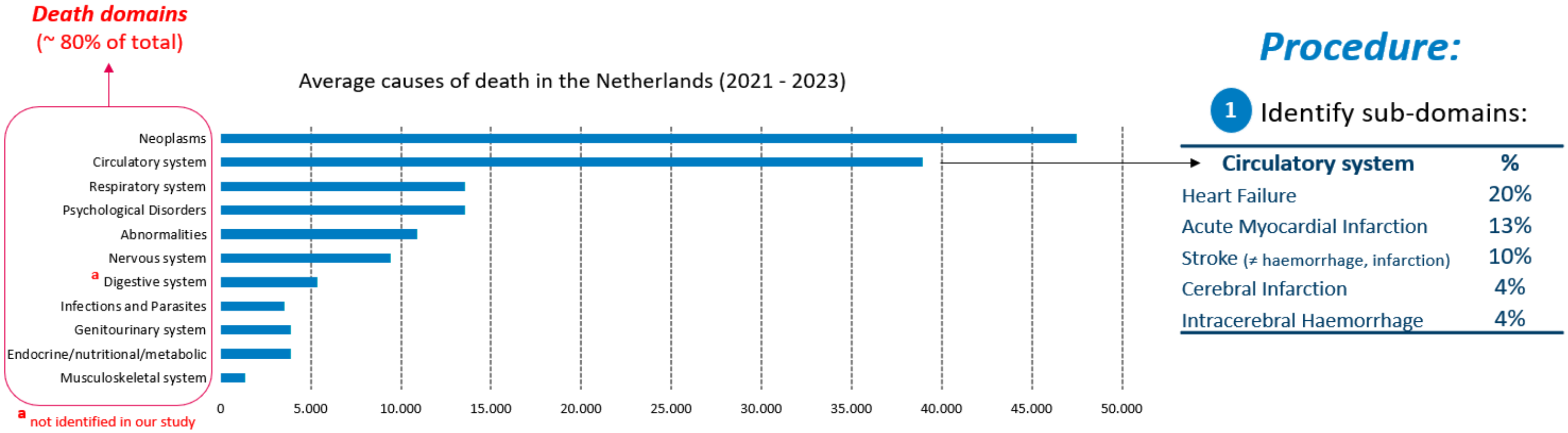


**Figure S8. | Procedure to identify specific diseases causes of death from diseases causes of death domains**
Figure exemplifies the procedure to fo from Disease Death Domains (right) to Specific Diseases Causes of Death. In the example we extract from the Circulatory System Domain the top 5 Specific Circulatory System Diseases
